# Can Severity of a Humanitarian Crisis be Quantified? Assessment of the INFORM Severity Index

**DOI:** 10.1101/2020.12.08.20246256

**Authors:** Velma K Lopez, Angeliki Nika, Curtis Blanton, Leisel Talley, Richard Garfield

## Abstract

**Background:** Those responding to humanitarian crises have an ethical imperative to respond most where the need is greatest. Metrics are used to estimate the severity of a given crisis. The INFORM Severity Index, one such metric, has become widely used to guide policy makers in humanitarian response decision making. The index, however, has not undergone critical statistical review. If imprecise or incorrect, the quality of decision making for humanitarian response will be affected. This analysis asks, how precise and how well does this index reflect the severity of conditions for people affected by disaster or war?

**Results:** The INFORM Severity Index is calculated from 35 publicly available indicators, which conceptually reflect the severity of each crisis. We used 172 unique global crises from the INFORM Severity Index database that occurred January 1 to November 30, 2019 or were ongoing by this date. We applied exploratory factor analysis (EFA) to determine common factors within the dataset. We then applied a second-order confirmatory factor analysis (CFA) to predict crisis severity as a latent construct. Model fit was assessed via chi-square goodness-of-fit statistic, Comparative Fit Index (CFI), Tucker-Lewis Index (TLI), and Root Mean Square Error of Approximation (RMSEA). The EFA models suggested a 3- or 4-factor solution, with 46% and 53% variance explained in each model, respectively. The final CFA was parsimonious, containing three factors comprised of 11 indicators, with reasonable model fit (Chi-squared=107, with 40 degrees of freedom, CFI=0.94, TLI=0.92, RMSEA=0.10). In the second-order CFA, the magnitude of standardized factor-loading on the ‘societal governance’ latent construct had the strongest association with the latent construct of ‘crisis severity’ (0.73), followed by the ‘humanitarian access/safety’ construct (0.56).

**Conclusions:** A metric of crisis-severity is a critical step towards improving humanitarian response, but only when it reflects real life conditions. Our work is a first step in refining an existing framework to better quantify crisis severity.

## Background

Humanitarian crises present a multitude of possible harmful health consequences for individuals (1). When populations are displaced, new settlements can have poor housing and sanitation conditions, leading to increased prevalence of acute infectious diseases, such as respiratory and enteric illnesses (2,3). Disruption of food systems can result in acute malnutrition (4,5) as well as further chronic malnutrition (6). In addition, interruption of health services limits the management of chronic diseases (7), access to sexual and reproductive health care (8), and distribution of immunizations among children(9). Mental health disorders, namely post-traumatic stress disorder, depression, and anxiety, are common among the displaced (10). While each of these morbidities are harmful on their own, they often interact, worsening overall wellbeing (1), and playing a role in the larger ecosystem that affects high mortality rates among crisis-affected people (11). Moreover, the impacts of humanitarian crises go beyond individual well-being to negatively influence communities, society, and the environment (12). Given the potentially devastating and longstanding impact of a humanitarian crisis, it is critical to provide aid where it is needed most.

The need for humanitarian assistance is great; yet there is limited funding available for response. For example, in 2020, the United Nations Office for the Coordination of Humanitarian Affairs (UNOCHA) estimated that there were 166.5 million people requiring humanitarian assistance and a gap of $14 billion (USD) in aid funding (13). With limited funding, it is imperative to assess which populations are in the greatest need and allocate resources accordingly. Currently, the United Nations (UN) system uses the crisis metric *number of people in need (PIN) of humanitarian assistance* to guide aid allocation. Because crises are diverse, the definition of PIN is non-specific (14) and the estimation is based on non-standardized data collection (15). Thus, using PIN as a basis of allocating resources may be limited. One step towards improving aid allocation is shifting the paradigm away from asking “how many people are in need?” and towards, “how bad are their needs?”. Accordingly, a systematic metric of crisis severity, that is “how bad is it?”, would reflect needs in ongoing crises and predict severity if conditions change (16). Applying a component measurement of crisis severity to aid allocation should help align resource distribution with core humanitarian principles: humanity, impartiality, independence, and neutrality.

Developing a metric to quantify the severity of a crisis is challenging. First and foremost, humanitarian crises are diverse and evolving events. Of the metrics that can be applied to a wide array of crises, most are designed for intra-country assessment of severity (e.g., the UNOCHA’s Humanitarian Needs Comparison Tool (17) or Kandeh et al.’s assessment of crisis-related vulnerability in Yemen (18)). While it is useful to assess geographic disparities, the need for humanitarian assistance is often based on estimates of overall crisis severity. For example, Bayram et al.’s 2012 Public Health Impact Severity Scale recommends using expert opinion to rank 12 indicators from the Sphere Project “Minimum Standards”, with the final severity score reflecting a weighted sum of the ranks (19). This framework, however, has yet to be implemented as the authors state limited availability of timely data. Eriksson et al. proposed a similar approach of ranking and summing key variables, but conceptualized severity as more holistic predictor of humanitarian need by drawing on psychological theory and ranked variable importance based on presence of the indicator in the literature (20). Like the Public Health Impact Severity Scale, use of their model has not been widespread.

The current model used to quantify crises severity is the INFORM Severity Index, a metric based on comparable data drawn from publicly available sources (21). Developed via partnerships and through consensus building among experts, the index uses a conceptual framework that describes crisis severity as a complex, multi-factorial construct after the immediate, emergency phase of crisis. The index is used by policymakers to set or justify priorities for providing humanitarian support, bring attention to unknown crises, and to monitor crisis trends. However, the model has not to date undergone statistical review.

We seek to critically evaluate the overall index model structure and assess the relationships between indicators. The INFORM Severity Index database, which was publicly available under the name ‘Global Crisis Severity Index (GCSI)’, included 172 unique crises in the version released in December 2019. These crises were either ongoing as of November 30, 2019, or had occurred earlier in 2019. The INFORM model inputs 35 unique indicators across three pillars (‘impact of the crisis’, ‘complexity of the crisis’, ‘conditions of the people’) to estimate crisis severity (see Table 1 for data and definitions and Appendix 1 for details regarding the GCSI construction). Because this is the most commonly used data source for humanitarian stakeholders to assess crisis severity, our objective is to determine if the entire model or a subset of its components could be used to estimate crisis severity through a score. Importantly, the GCSI dataset tracks diverse crises and analysis of it provides insight into severity of a wide range of emergency events. Here, we applied factor analysis, a method commonly used for data reduction of highly correlated and grouped data. This review is an attempt to generate a more robust estimate of crisis severity.

**Table 1.**
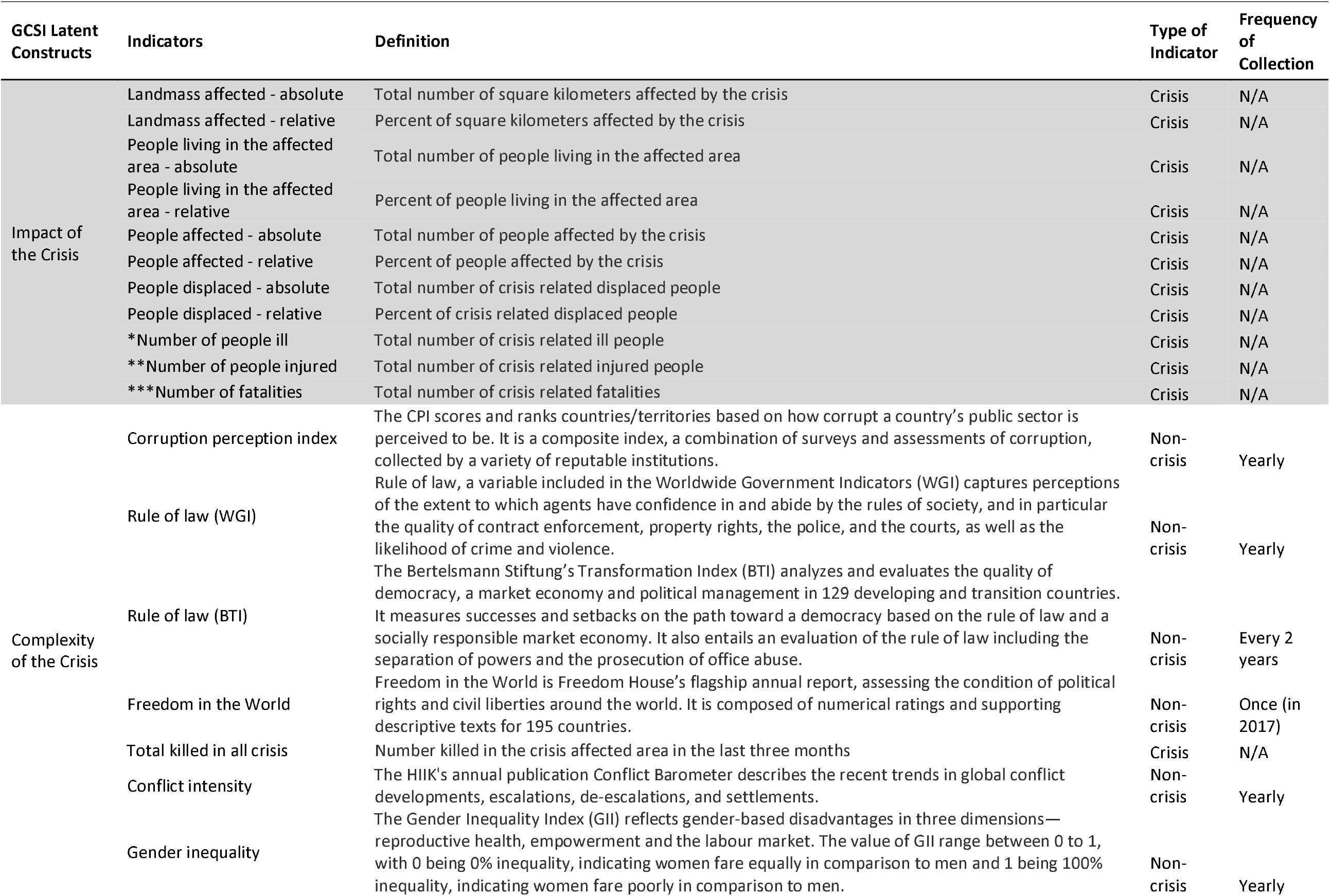
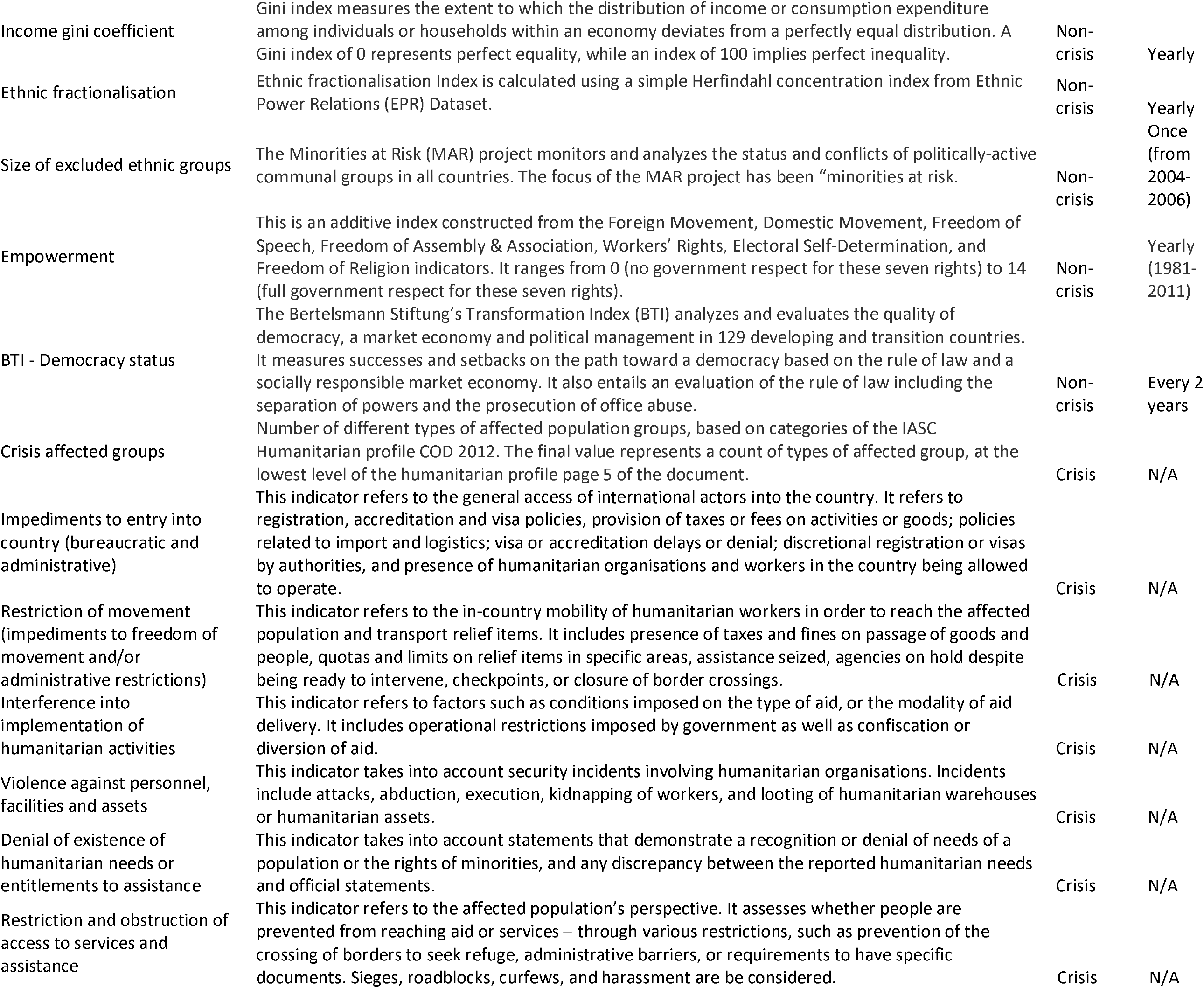
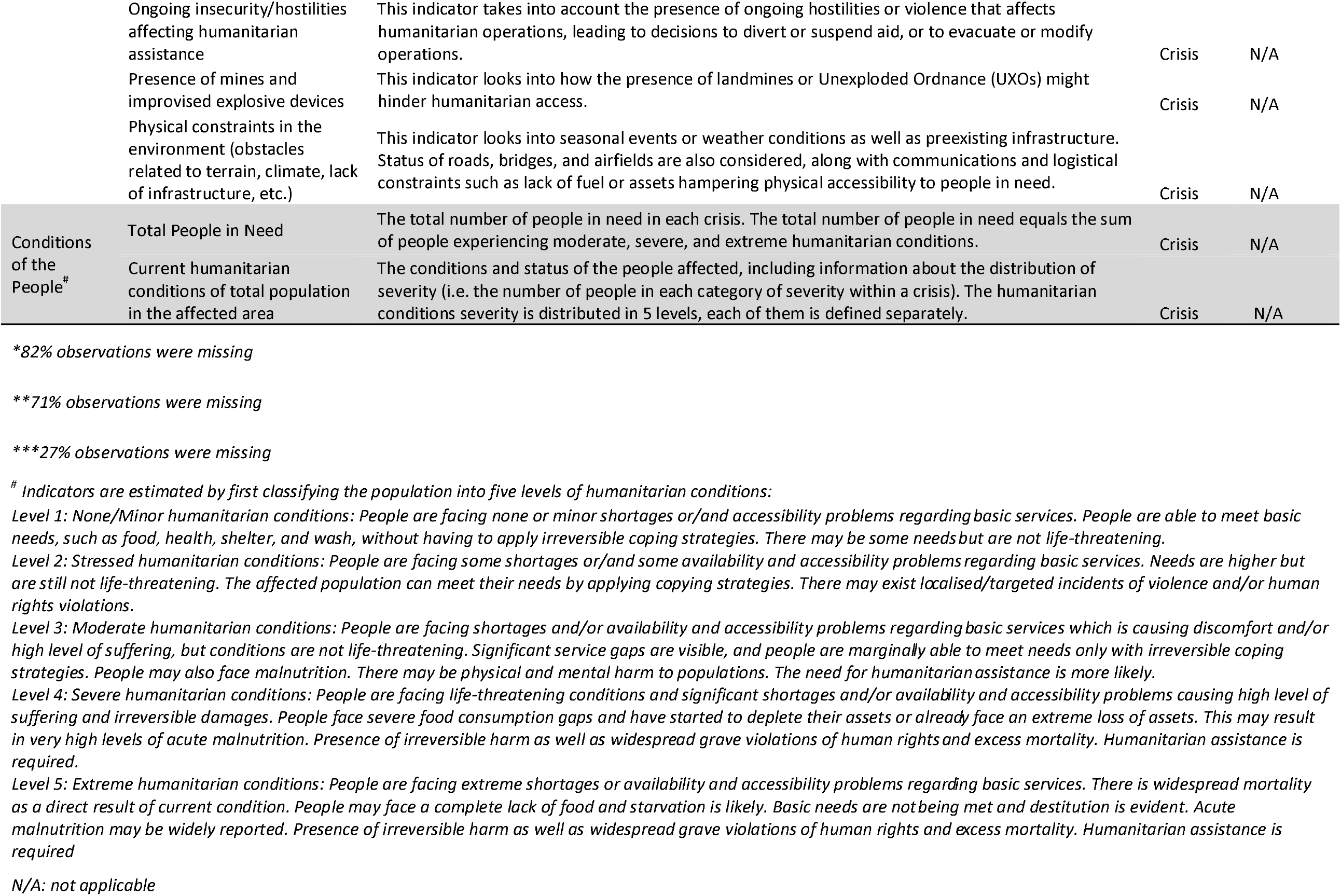
Global Crisis Severity Index (GCSI) pillar, which we consider latent constructs, presented alongside the indicators within each construct, definition, whether the indicator is crisis related, and if not, when the data are routinely collected. All indicators, and their references, are further described within the GCSI spreadsheet under the sheet titled “Indicator Metadata”: https://data.humdata.org/dataset/inform-global-crisis-severity-index.

## Methods

### Data

We analyzed data from the beta version of the INFORM Severity Index database, which was publicly available under the name ‘Global Crisis Severity Index (GCSI)’ and released in December 2019 (33). We extracted data from 172 unique global crises that were either ongoing as of November 30, 2019, or had occurred earlier in 2019. Appendix 1 describes how the INFORM Severity Index is calculated.

### Measures

The GCSI uses a total of 35 ordinal indicators to represent three pillars (impact of the crisis, complexity of the crisis, conditions of the people), which we consider latent constructs (Table 1). Each ordinal indicator is scored based on continuous variables. The first construct, ‘the impact of the crisis’, is comprised of 11 indicators, all of which are ordinal versions of data collected from the specific crisis. The second construct, ‘the complexity of a crisis’, is comprised of 22 indicators. Of these indicators, 12 are publicly available indices; one is an ordinal version of data collected from the specific crisis; and the remaining nine indicators reflect qualitative information that is given a quantitative score. The final construct, ‘conditions of the people’, has two indicators, each of which uses estimates of the number of people in need of humanitarian assistance for the given crisis. Spearman’s rank correlation coefficients for all indicators are presented in Appendix 1. From the 35 GCSI indicators, we removed three indicators that had more than 25% of observations missing.

### Analytical Approaches

Modeling a construct such as ‘severity’ requires leveraging information from multiple indicators. Any approach that does not account for correlation between the indicators will likely result in imprecise final estimates. Thus, our analytical framework uses structural equation modeling to predict crisis severity. This method explicitly includes measurement error for each indicator, assessment of model fit – both overall and at the indicator level -, and prediction from optimal combinations of indicators. The overall goal of the analysis is to deduce causal relationships by accounting for correlation coefficients. To do so, we have a two-step approach. First, we used exploratory factor analysis (EFA) to identify patterns of grouping among the indicators. This step provided insight on whether the data supported the GCSI pillar construction. We then applied confirmatory factor analysis (CFA) to test whether the identified relationships from the EFA were statistically meaningful and if the multiple latent constructs were inter-related (as hypothesized by the GCSI pillar construction).

#### Exploratory Factor Analysis

We evaluated the relationships between 32 indicators in the GCSI conceptual framework through EFA. Based on an initial scree plot, we employed four maximum likelihood EFA models, ranging from 3- to 6 -factor solutions, each with an oblimin rotation, that is, correlation was permitted between factors (34). Missing values were imputed with the indicator median within all EFAs. We evaluated the models for the following characteristics: sums of squared loadings greater than 1.0 for each factor; factors that contribute to at least 10% to the overall variance; and collective contribution of at least 60% of the overall variance. Next, we reviewed the indicator factor loadings to identify latent constructs within the dataset.

Using the information learned from the EFA models, we removed indicators from the dataset if they did not provide unique information to identified factors as their inclusion in a final score could lead to either bias or imprecision. Indicators were removed if they had factor loadings less than 0.30 or cross-loaded onto more than one factor with a loading less than 0.20, or if cross-loadings had values in opposite directions (for example, 0.37 and -0.33).

#### Confirmatory Factor Analysis

With the reduced dataset and using standardized indicators, we applied a full information maximum likelihood CFA to model crisis severity. First, we built first-order CFAs with relationships identified in the EFA. We removed indicators from the CFAs if they had residuals greater than 0.10 with indicators on different latent constructs. We also added covariances between indicators on the same latent construct if their residual correlation was greater than 0.10. Finally, we added a second-order latent construct to the model, which represented ‘crisis severity’. Model fit was assessed via chi-square goodness-of-fit statistic, Comparative Fit Index (CFI), Tucker-Lewis Index (TLI), and Root Mean Square Error of Approximation (RMSEA). Acceptable model fit was evaluated using recommended cut-offs characterized as CFI and TLI greater than 0.90 and RMSEA less than 0.08 (35).

We also estimated values for the latent crisis severity variable (i.e., factor scores) based on the factor loadings in the second-order CFA. Latent severity scores were normalized to range from zero to one.

We conducted several subanalyses to determine the robustness of the overall results. These analyses focused on: incorporating the ‘people in need of humanitarian assistance’ indicator within the final models (see Appendix 5); data quality implications (see Appendix 6); comparison of the modeled scores to the original scores (see Appendix 7); the role of governance when estimating crisis severity (Appendix 8); the implication of missing data (see Appendix 9); and the overall model fit bootstrapped subsamples of the dataset (see Appendix 10). Analyses were conducted using the R (version 3.6.2) packages *psych, GPArotation*, and *lavvan*; see Appendix 4 for primary analyses’ R code.

The Human Research Protection Office within the Center for Global Health at the Centers for Disease Control and Prevention reviewed the study and determined it to be non-research.

## Results

### Original GCSI Score

The GCSI includes a large number of indicators (Table 1), which reflect both crisis related data and non-crisis related data. The original GCSI score is generated from combining these indicators into pillars, which are then aggregated into a score. For example, the complex emergency in Somalia, coded as *SOM001* in the GCSI database, was classified as having “High Severity” with a score of 4.0 as of November 2019. Qualitatively, there is concurrence between the GCSI score given to the Somali crisis and the nation’s social structure and events that have occurred there: approximately four million people needed humanitarian assistance in Somalia in 2019, and millions had been displaced by recurring conflict, insecurity, forced evictions, drought, and floods (see https://www.unocha.org/somalia). The GCSI Severity Score for Somalia was estimated via a weighted mean of the values derived for each pillar: 4.4 (with a 0.2 weight applied) for the Impact of the Crisis, 4.4 (with a 0.3 weight applied) for the Complexity of the Crisis, and 3.0 (with a 0.5 weight applied) for the Conditions of the People. The data feeding into these estimates, and their values, are presented in Figure 1. Within the figure, each box is a data point, each oval represents the aggregation of the boxes (or other ovals) preceding it (represented by an arrow), and each circle represents the aggregation into the GCSI pillars. Importantly, each pillar calculation, and the calculation of the sub-indicators used in the pillar score, is unique (see Appendix 1 for details). Briefly, the pillar scores are derived using the following approaches:

**Figure 1.**
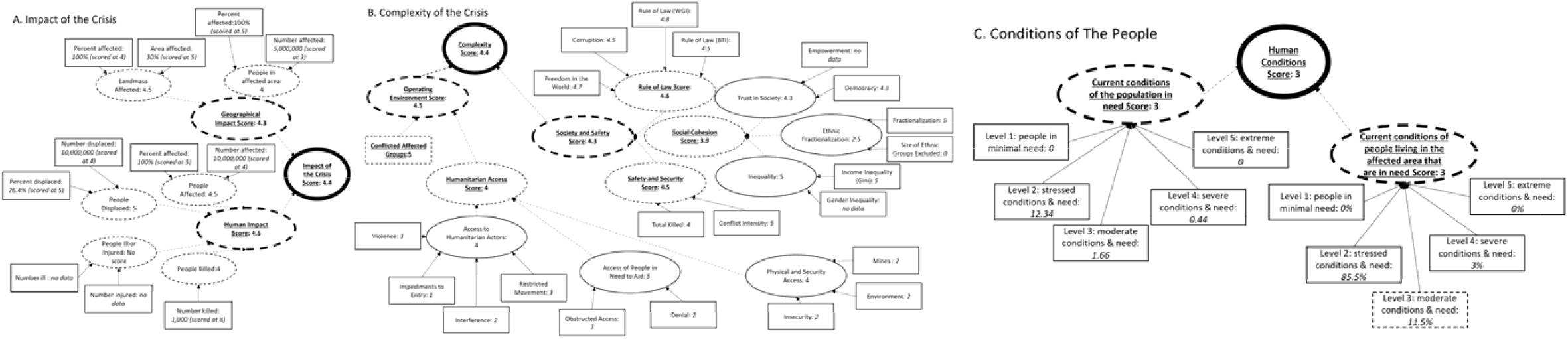
A schematic of the GCSI conceptual framework for the complex crisis in Somalia (coded as *SOM001* in the GCSI database). Each box represents a data point, each oval represents the aggregation of the boxes (or other ovals) preceding it (represented by an arrow), and each circle represents the aggregation into the GCSI pillars. Shapes with dashed values represented aggregated scores of sib-indicators and bold shapes are the aggregated final scores. Panel A shows the Impact of the Crisis. Panel B shows the Complexity of the Crisis. Panel C shows the Conditions of the People. In panel C, the Condition of the population in Need Score shows values that are scaled to 1,000,0000 people.

⍰ <u>The Impact of the Crisis (Figure 1A)</u> is the weighted sum of two composite indicators - The Human Impact (weighted at 0.7) and the Geographical Impact (weighted at 0.3). The Human Impact Score is an aggregate of 4 sub-indicators, however, for Somalia, data are only available for 3 components. The Geographical Impact is a mean score generated from two sub-indicators.
⍰ <u>The Complexity of the Crisis (Figure 1B)</u> is estimated by calculating the geometric mean of two composite indicators – Society and Safety and Operating Environment. The Society and Safety Score reflect a mean of three sub-indicators, each with a different number of data inputs. Missing inputs, such as ‘Gender Inequity’ in Somalia, are ignored during aggregation. The Operating Environment Score is estimated from the average value of a sub-indictor and a data input variable. However, here, the sub-indicator aggregation mostly reflects summation, and is scaled if there are more than two variables that contribute to the sub-indicator.
⍰ <u>Conditions of the People as a Result of the Crisis (Figure 1C)</u> is the average of two sub-indicators – Current Humanitarian Conditions of the Total Population, and Current Humanitarian Conditions of the Population Affected. Here, the population is ranked into one of five levels: 1. those facing minimal humanitarian need, 2. those in stressed humanitarian conditions and needs, 3. those in moderate humanitarian conditions and needs; 4. those in severe humanitarian conditions and needs, and 5. those in extreme humanitarian conditions and needs. The Current Humanitarian Conditions of the Total Population sub-indicator reflects the sum of people in levels 3-5, which is then ranked. The Current Humanitarian Conditions of the Population Affected sub-indictor, however, is calculated slightly differently. Here, the highest level is taken if the percent of the population affected at that level is greater than 5%. For example, the Somalia crisis is given a value of 3 for this indicator because 11.5% of the population affected fall into level 3 of need.

Overall, Figure 1 paints an intricate, and convoluted, picture of data relationships used to classify the severity of the Somali crisis. Understanding these relationships sheds insight into how these data can be used to generate severity values.

### GCSI Analysis

To guide our analysis, we first examined the following characteristics of the GCSI data frame: correlation between indicators, distributions of indicators, and the proportion of non-missing data. The indicators are highly correlated (see Appendix 1) within the conceptual GCSI pillars and between them, with approximately 31% of the indicators having correlation coefficients greater than +/-0.6. The correlation values suggest complex underlying relationships, and inference thereof required a method that accounts for statistical dependencies. Mean and median values of the ordinal scores did not differ greatly for most indicators, suggesting only slightly skewed distributions (Table 2) and the appropriate application of parametric methods. Three indicators had a substantial proportion of missing data (‘Number of people ill’; ‘Number of people injured’; ‘Number of fatalities’), so we removed them from our analysis. Seventy-five to 100% of observations were available for the remaining 32 indicators. The indicators related to people displaced from a crisis had the two lowest number of observations (75% and 77% of total observations), as did the indicator for people in need (80% of total observations).

**Table 2.**
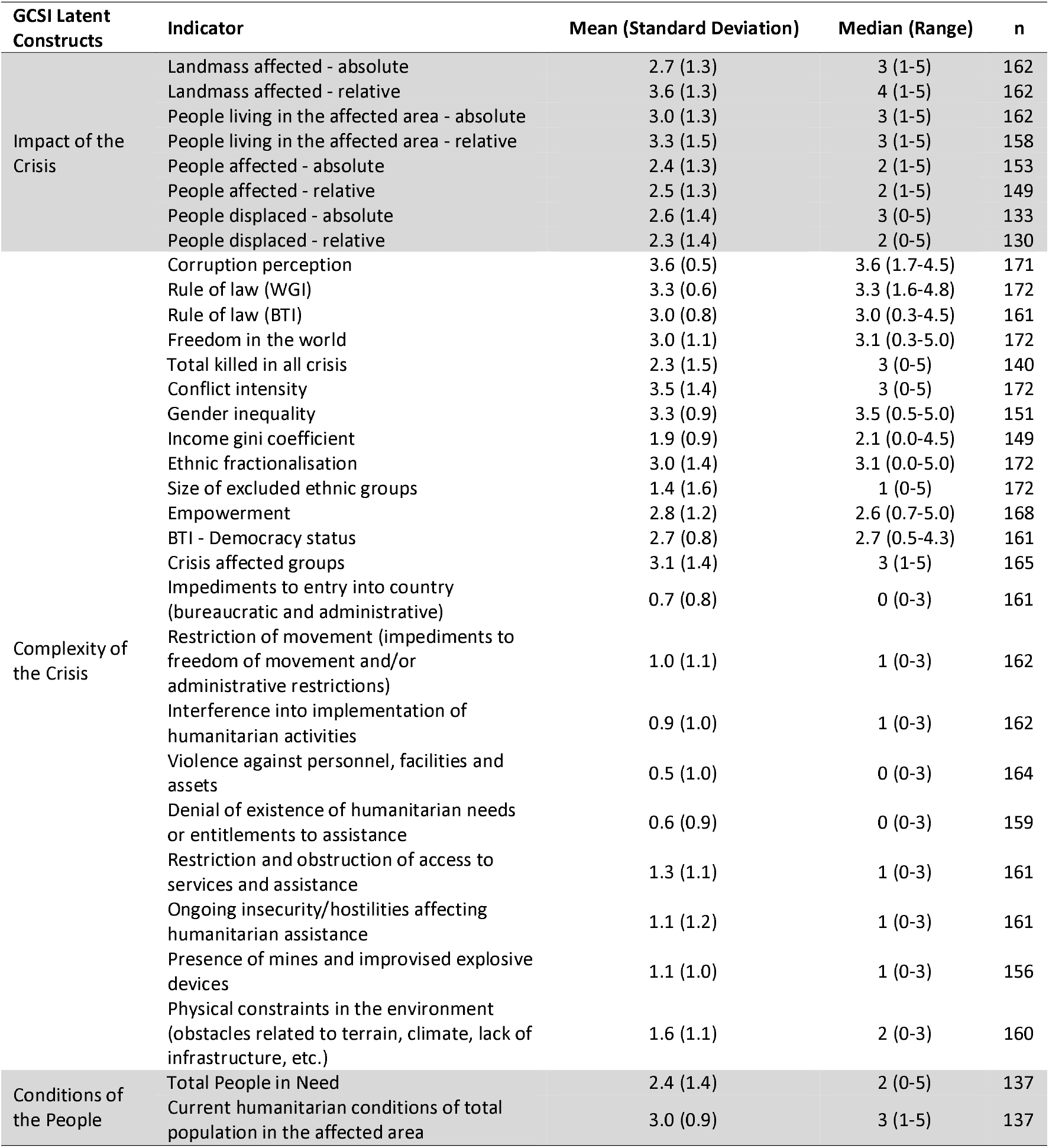
Descriptive statistics for Global Crisis Severity Index (GCSI) indicators: Mean, standard deviation, median, range, and total number of observations (n) are presented for each indicator. Indicators are highlighted to reflect GCSI latent constructs (*GCSI dataset, 2019, N=172*).

We applied factor analysis to test whether the indicators in the GCSI dataset could be used to generate an estimate of crisis severity. Our approach had two primary steps: exploratory factor analysis (EFA), followed by confirmatory factor analysis (CFA). We removed all “pillar” aggregated values and only used the input data to generate our models. In the EFA model, we imputed median values of a given indicator if the observation was missing, while in the CFA model, we used maximum likelihood to address missing information. To assess whether these approaches influenced the final model estimates, we also ran the EFA model using case deletion for missing observations and the CFA model using multiple imputation (Appendix 9). We found negligible differences between the results from these different approaches and those presented here (Appendix 9).

#### EFA

We examined 32 of the 35 GCSI variables to assess their grouped correlation patterns. The EFA models suggested a 3- or 4-factor solution (Table 3). While the 5- and 6-factor solutions had greater cumulative variance explained. The proportion variance explained for each factor did not add substantive information to the model. This was also evident in the indicator factor loadings for these models, which showed more cross-loadings between indicators on factors with less than 10% proportion variance explained (see Appendix 2 for factor loadings for 5- and 6-factor solutions).

**Table 3.** Exploratory Factor Analysis summary information for each factor solution identified: sums of squared loadings, proportion variance explained and cumulative variance (*GCSI dataset, 2019, N=172*).

| Factors | Three-Factor Solution |  |  | Four-Factor Solution |  |  | Five-Factor Solution |  |  | Six-Factor Solution |  |  |
| --- | --- | --- | --- | --- | --- | --- | --- | --- | --- | --- | --- | --- |
|  | *Sums of Squared Loadings* | *Proportion Variance Explained* | *Cumulative Variance Explained* | *Sums of Squared Loadings* | *Proportion Variance Explained* | *Cumulative Variance Explained* | *Sums of Squared Loadings* | *Proportion Variance Explained* | *Cumulative Variance Explained* | *Sums of Squared Loadings* | *Proportion Variance Explained* | *Cumulative Variance Explained* |
| Factor 1 | 4.76 | 0.15 | 0.15 | 4.55 | 0.14 | 0.14 | 2.49 | 0.08 | 0.08 | 4.6 | 0.14 | 0.14 |
| Factor 2 | 3.67 | 0.11 | 0.26 | 3.81 | 0.12 | 0.26 | 4.66 | 0.15 | 0.23 | 2.45 | 0.08 | 0.22 |
| Factor 3 | 6.56 | 0.20 | 0.46 | 6.58 | 0.21 | 0.47 | 3.37 | 0.11 | 0.34 | 5.57 | 0.18 | 0.4 |
| Factor 4 | - | - | - | 1.97 | 0.06 | 0.53 | 5.96 | 0.19 | 0.53 | 1.84 | 0.06 | 0.46 |
| Factor 5 | - | - | - | - | - | - | 1.97 | 0.06 | 0.59 | 3.18 | 0.10 | 0.56 |
| Factor 6 | - | - | - | - | - | - | - | - | - | 2.02 | 0.06 | 0.62 |

Additional examination of the factor loadings in 3- and 4-factor models highlighted three primary findings (Table 4). First, several indicators had factor loadings less than 0.30, which implies that they do not contribute to any of the factors. Second, indicator cross-loadings onto multiple factors were common, and thus, these indicators did not provide unique information. Finally, the indicators grouped into a pattern similar to sections of the GCSI conceptual framework. In both solutions, factor 1 was comprised of indicators related to societal constructs (and originally conceptualized as part of the ‘complexity of the crisis’), while indicators within the ‘impact of the crisis’ construct grouped together in factor 2. Factor 3 was comprised of indicators related to humanitarian access and safety; while the fourth factor was a further disaggregation of factor 2. Of note, the EFA results did not show that indicators related to ‘conditions of the people’ had mathematical importance. Indicators excluded from subsequent CFA models are shown in Table 4.

**Table 4.** Global Crisis Severity Index (GCSI) Indicators factor loadings for each Exploratory Factor Analysis (EFA) factor solution. Factor loadings are only presented if greater than the absolute value of 0.3 (*GCSI dataset, 2019, N=172*).

| Indicators | Three-Factor Solution |  |  | Four-Factor Solution |  |  |  |
| --- | --- | --- | --- | --- | --- | --- | --- |
|  | Factor 1 | Factor 2 | Factor 3 | Factor 1 | Factor 2 | Factor 3 | Factor 4 |
| *Landmass affected - absolute |  | 0.43 | 0.32 |  | 0.45 | 0.32 |  |
| Landmass affected - relative |  | 0.86 |  |  | 0.86 |  |  |
| *People living in the affected area - absolute |  | 0.49 | 0.37 |  | 0.49 | 0.33 |  |
| People living in the affected area - relative |  | 0.93 |  |  | 0.93 |  |  |
| *People affected - absolute |  | 0.51 | 0.38 |  | 0.53 | 0.35 |  |
| People affected - relative |  | 0.74 |  |  | 0.75 |  |  |
| **People displaced - absolute |  |  | 0.49 |  |  | 0.44 | -0.41 |
| **People displaced - relative |  | 0.36 |  |  | 0.37 |  | -0.33 |
| Corruption perception | 0.64 |  |  | 0.51 |  |  |  |
| *Rule of law (WGI) | 0.56 |  | 0.35 | 0.42 |  | 0.42 | 0.47 |
| Rule of law (BTI) | 0.96 |  |  | 0.96 |  |  | 0.49 |
| Freedom in the world | 0.89 |  |  | 0.87 |  |  |  |
| Total killed in all crisis |  |  | 0.72 |  |  | 0.73 |  |
| Conflict intensity |  |  | 0.70 |  |  | 0.71 |  |
| Gender inequality | 0.38 |  |  |  |  |  | 0.51 |
| Income gini coefficient |  |  | -0.35 |  |  | -0.31 | 0.41 |
| *Ethnic fractionalisation |  |  |  |  |  |  |  |
| *Size of excluded ethnic groups |  |  |  | 0.34 |  |  | -0.31 |
| Empowerment | 0.47 |  |  | 0.51 |  |  |  |
| BTI - Democracy status | 0.93 |  |  | 0.95 |  |  |  |
| Crisis affected groups |  |  | 0.54 |  |  | 0.54 |  |
| *Impediments to entry into country (bureaucratic and administrative) | 0.36 |  | 0.35 | 0.46 |  | 0.31 | -0.34 |
| Restriction of movement (impediments to freedom of movement and/or administrative restrictions) |  |  | 0.87 |  |  | 0.88 |  |
| Interference into implementation of humanitarian activities |  |  | 0.67 |  |  | 0.65 |  |
| Violence against personnel, facilities and assets |  |  | 0.57 |  |  | 0.58 |  |
| Denial of existence of humanitarian needs or entitlements to assistance |  |  | 0.31 | 0.35 |  |  | -0.39 |
| Restriction and obstruction of access to services and assistance |  |  | 0.80 |  |  | 0.79 |  |
| Ongoing insecurity/hostilities affecting humanitarian assistance |  |  | 0.76 |  |  | 0.79 |  |
| Presence of mines and improvised explosive devices |  |  | 0.62 |  |  | 0.64 |  |
| Physical constraints in the environment (obstacles related to terrain, climate, lack of infrastructure, etc.) | 0.38 |  |  |  |  |  | 0.34 |
| *Total People in Need |  | 0.51 | 0.40 |  | 0.53 | 0.36 |  |
| *Current humanitarian conditions of total population in the affected area |  |  |  |  |  |  |  |

\*Indicator excluded from subsequent CFA models
\*\*Indicator excluded from CFA model with 4 latent constructs, but included in CFA with 3 latent constructs

#### CFA

We initially built four different CFA models to reflect the relationships identified with the factor loadings in the EFAs; each of the four models had an increasing number of latent constructs (from three constructs to six constructs).

The CFA with three latent constructs (base model) was appropriately specified but showed poor fit (Table 5). Indicators were removed and covariances added to reflect the residual correlations of indicators across the dataset (see Appendix 3 for correlation matrix) until the best model fit was generated (final model in Table 5). The final CFA contained 11 indicators: rule of law, democracy, freedom, gender inequality, empowerment, number of people killed, restricted movement, obstructed access to assistance, percent of landmass affected, people living in the affected area, people affected. We used this model to create a second-order CFA (Figure 2). The model fit statistics of the second-order CFA were the same as the fit statistics of the final first-order CFA model (Table 5). In the second-order CFA, the magnitude of standardized factor-loading on the ‘societal governance’ latent construct had the strongest association with the latent construct of ‘crisis severity’ (0.73), followed by the ‘humanitarian access/safety’ construct (0.56).

**Table 5.**
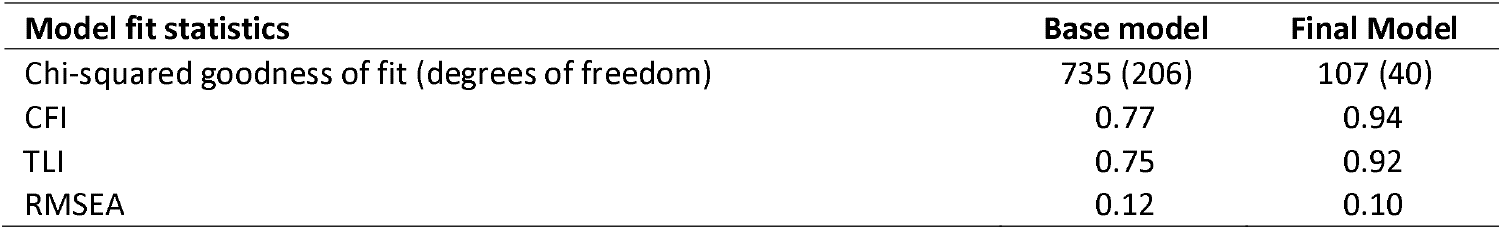
Fit statistics for first-order Confirmatory Factor Analysis (CFA) models: Chi-squared goodness of fit test statistic, Comparative Fit Index (CFI), Tucker-Lewis Index (TLI), and Root Mean Square Error of Approximation (RMSEA) Index (*GCSI dataset, 2019, N=172*).

| **Model fit statistics** | **Base model** | **Final Model** |
| --- | --- | --- |
| Chi-squared goodness of fit (degrees of freedom) | 735 (206) | 107 (40) |
| CFI | 0.77 | 0.94 |
| TLI | 0.75 | 0.92 |
| RMSEA | 0.12 | 0.10 |

**Figure 2.**
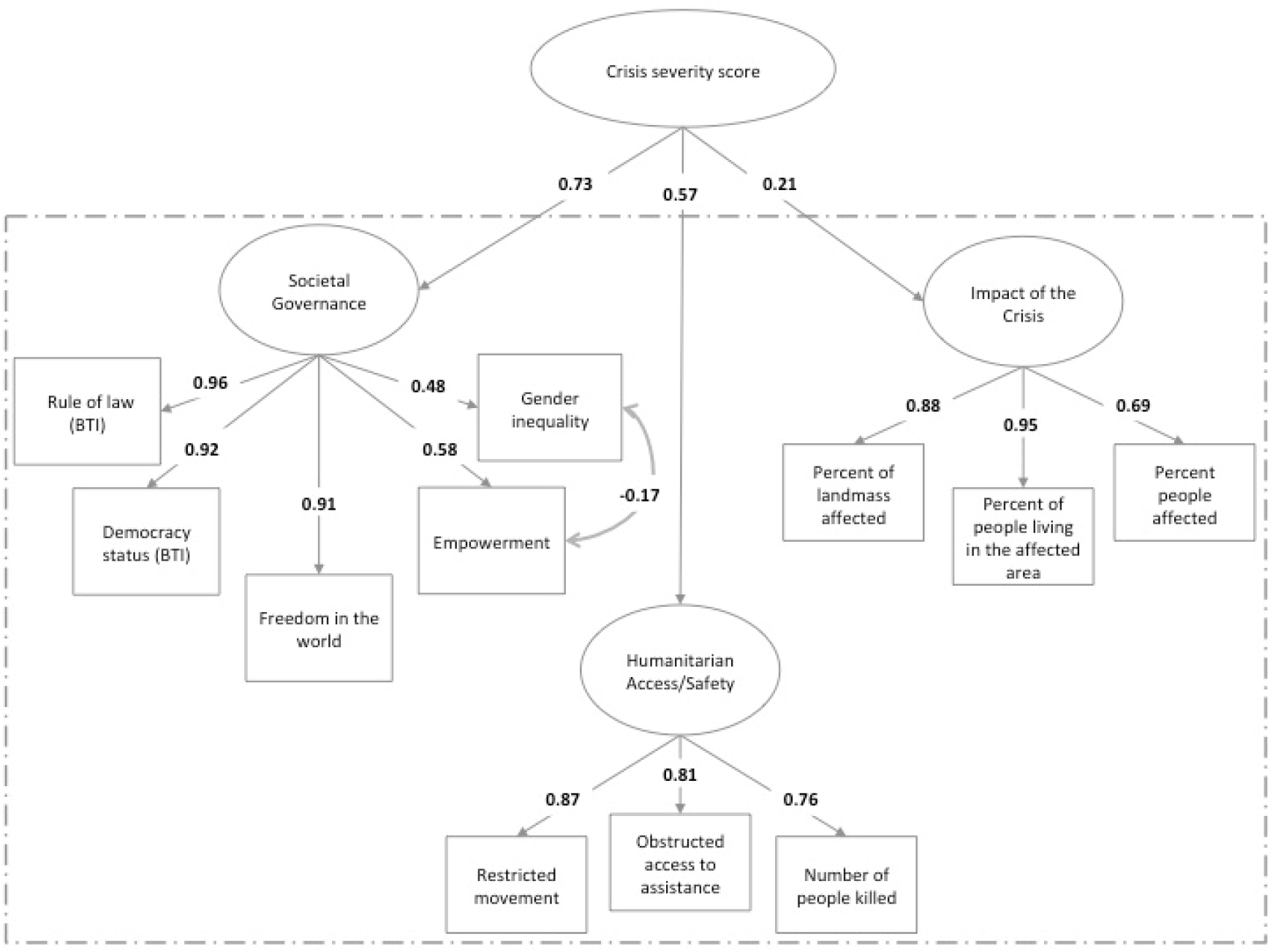
Second-order Confirmatory Factor Analysis (CFA), with factor loadings. The ovals reflect latent variables and the boxes reflect indicators. The dashed box contains the final first-order CFA.

The CFA with four latent constructs had a non-positive covariance matrix when a second order latent variable was added. No solutions were found for the 5 or 6 latent variable models.

#### Severity score

We used the final CFA model to generate normalized severity scores for each crisis (which range between 0 and 1 to represent low to high severity). The mean and median latent severity score for all crises were similar, at 0.53 and 0.54, respectively. Severity scores were highest in complex crises and fell within the upper two-thirds of all scores (Figure 3). Regional crises, conversely, had a lower mean severity score. These types of crises fell into the bottom two-thirds of the range. Crises in countries that had a mean severity score of greater than 0.90 included Syria, Somalia, Yemen, and The Democratic People’s Republic of Korea, whereas countries with mean severity scores less than 0.10 included Costa Rica and Brazil.

**Figure 3.**
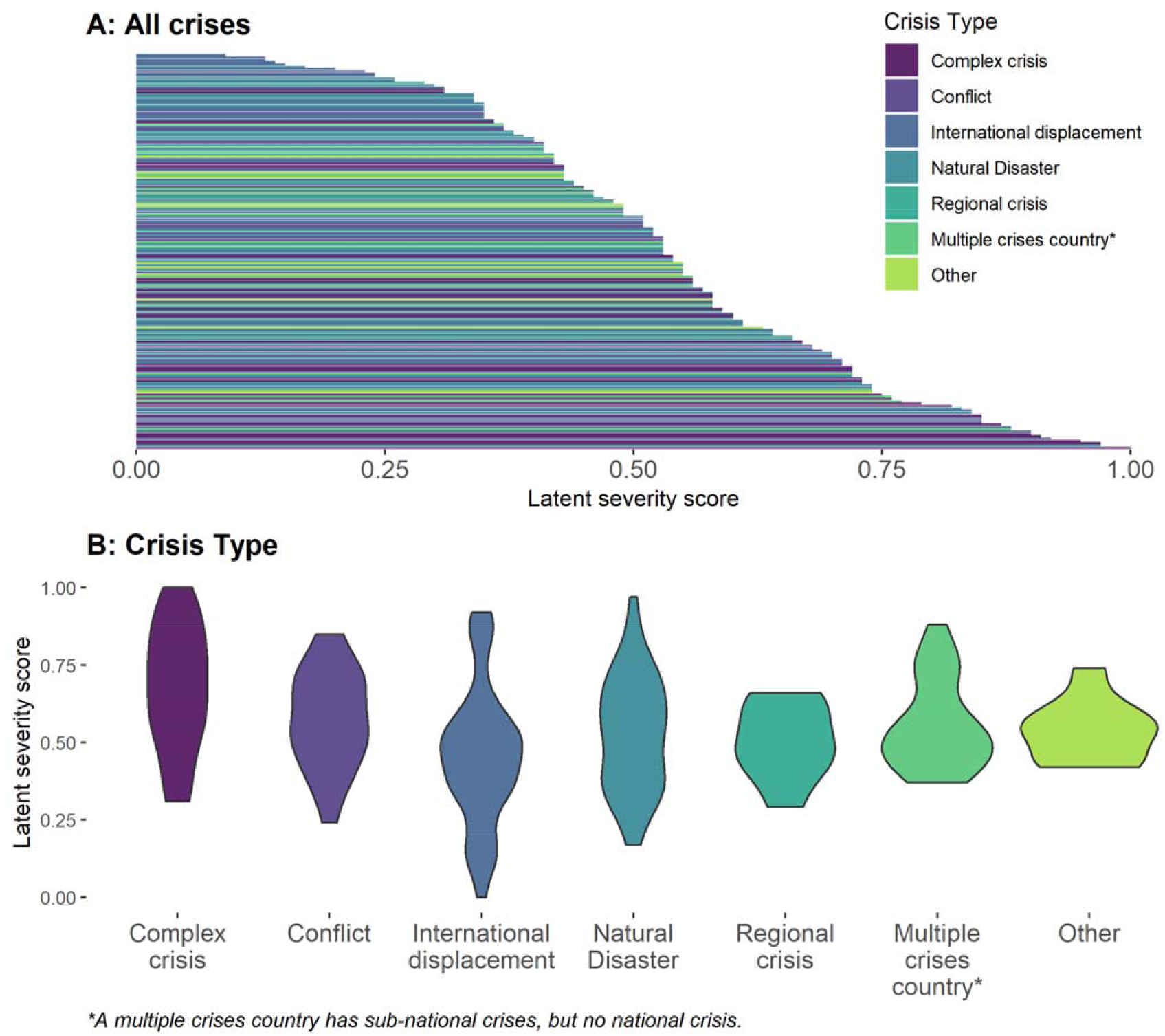
The distribution of latent crisis severity scores. Figure 2A shows all crises (n=172) and Figure 2B is stratified by crisis type.

## Discussion

Our analyses show that crisis severity can be measured best through use of only 11 of the 35 GCSI indicators. Put another way, 24 of the 35 indicators used in the GCSI model do not contribute useful numerical information. In our final model, the strongest predictors of severity were a suite of indicators related to social structure/governance of a given nation state (rule of law, freedom, gender inequality, and empowerment), followed by indicators that were proxy measurements of humanitarian access/safety (number of people killed, restricted movement, and obstructed access to assistance). Weaker, although still relevant, predictors were related to the crisis impact on people and the environment. Overall, this analysis suggests that most of the key variables to estimate the severity of humanitarian need can be assessed globally, can be collected in a comparable way from one country to another, and do not depend on sudden changes to local conditions that would be unavailable to those making the calculations. In short, despite changing conditions and limited or imperfect information, we can use existing data sources to make reasonable estimates of severity around the world. Refinements to the existing GCSI model will make it easier and more reliable to make these estimates.

Holistically, the 11 selected indicators suggest that fragile states with limited accessibility for humanitarian actors have worse humanitarian conditions. This final model aligns with humanitarian actors’ experiences. Indeed, good governance is intrinsically related to avoiding or mitigating a humanitarian crisis (12). We tested the role of governance in our models (see Appendix 8) and found it to be a key latent constructure of severity, but only when crisis related variables were also included in the model. Broadly, economic and political stability are key components to this success, with inequality between social groups cited as a driver of crises and conflicts (22). It is unsurprising that humanitarian practitioners call for more robust inclusion of conflict early warning into preparedness systems for humanitarian crises (23). Indeed, considerable funding has been provided to post-conflict states for democracy development and peacebuilding, albeit with mixed success (24,25). Ample evidence supports these patterns, as data from the last 15 years show most humanitarian crises are re-occurring in the same countries, many of which are fragile states (13). Chad, the Central African Republic, the Democratic Republic of Congo, Somalia, and Sudan have all had 15 crises between 2005 and 2015.

Beyond governance, access to reach those in need is also important to reducing crisis severity. Humanitarian access, the ability to reach the most vulnerable, can be limited through various mechanisms. Restricted movements, which are common in conflicts and complex humanitarian crises, inhibit connections between aid workers and communities (26). Access can also be reduced through violence and obstruction. Within armed conflicts, bureaucratic and security constraints, and violence against aid workers and facilities distributing aid, have been cited as rationale for greatly reduced humanitarian access (26,27). For example, in the Syrian crisis, which is considered one of the worst in the world by humanitarian experts, UNOCHA reported that 1.1 million people were in need of humanitarian assistance in hard-to-reach-places in 2018; during this same year, access was inhibited by 142 attacks on health facilities, with 102 people dead and 189 injured (28). Thus, it is not surprising that our model results give weight to indicators reflecting the quality of humanitarian access (e.g., restricted movements, obstructed access, and number of people killed) for a given crisis.

Importantly, our final model differs from the original GCSI in two fundamental ways. First, we presented a parsimonious model, which removed 24 GCSI indicators. Using the reduced set of 11 indicators, the model showed acceptable fit, but had slightly higher error than the standard cut points; however, some debate exists on the usefulness of applying a single heuristic to assess model fit within factor analysis (29). The original GCSI was calculated using inconsistent approaches, and notably, does not account for basic statistical properties of correlated data. The high correlation in the dataset inhibits meaningful interpretation of combined values from the indicators. Second, we removed an entire GCSI pillar (‘conditions of the people’) as a result of insights from the EFA models, which has programmatic significance. Indeed, the data underlying the excluded indicators are routinely collected to estimate the number of people in need of humanitarian assistance. Given the strong value of these indicators to practitioners, we re-ran the final model and included these two indicators as standalone independent variables (Appendix 5). Of note, we did not include the two indicators as latent constructs, as our EFA analyses showed that they were not correlated. This sensitivity analysis suggested that a model including the *number of people affected* indicator has comparable model fit and yields similar severity scores to the second-order CFA model.

Our analysis, however, is limited by the data available for inclusion. First, the index includes a combination of static and dynamic variables. It is possible that static variables, such as those used to estimate social structure/governance are distal determinants of a crisis, rather than proximal measures. Additionally, we used population average data, which masks any disparities experienced within a population. Several population groups, namely, children, women, and the disabled, have worse crisis-related health outcomes than the rest of the population. Moreover, data from humanitarian crises are difficult to obtain, highly inaccurate, and highly correlated. While our sensitivity analyses assessing data quality suggest that our final model contained data that was no more or less reliable than the indicators excluded (Appendix 6), we cannot account for the lack of precision within the dataset. We included two indicators based on expert assessment of qualitative information (restricted movement, and obstructed access to assistance), which may be subject to imprecision or bias. Likewise, mortality estimates, which we also included in the final model, have been contested for accuracy in past crises (30,31). Additionally, the indicator for ‘relative people living in the affected area’ is highly correlated with many of the other variables in the final model. In an ideal scenario, this indicator would be removed from the model, however, when it was, the models did not converge. Thus, one limitation of retaining the variable is a slightly higher error than desired. Finally, we are limited in our ability to test the generalizability of the model given the small sample size and lack of additional data for testing. Nevertheless, our comparison of the model fit statistics and factor loadings to suggest that the model performance is consistent and unlikely overfit to the data (Appendix 10).

Importantly, a gold standard for crisis severity is unavailable to validate our model results and out-of-sample data were not available to assess predictions. In lieu of traditional validation, we compared the latent severity scores to the original GCSI scores (Appendix 7). These robustness checks suggested that the latent severity score may be a closer measure to true crisis severity than the original GCSI. Despite the limitations with data availability and independent data source for validation, we emphasize that this work is a first step towards improving crisis severity measurement. Because the calculations are derived from a model that weights indicators based on their correlations, estimating severity for a new crisis would require re-running the final CFA. Further research is needed to assess the feasibility of linking this framework with a field friendly application for humanitarian actors after additional analyses have been conducted.

## Conclusion

UN-coordinated humanitarian responses are lasting longer (32), with the average 2005 response ongoing for about four years compared to the 2017 response of seven years. Meanwhile, human and financial resources for humanitarian response are limited. More complicated responses, coupled with calls for increased resources, emphasize the need for objective tools to guide resource allocation. Indeed, a metric of crisis severity can add powerful contributions to determine priorities for humanitarian response, highlighting whether severity and subsequent aid/response align. However, a metric of crisis-is only useful if the metric is scientifically robust. Our work is a first step in refining an existing framework to quantify crisis severity. We suggest three additional areas of needed exploration. As presented here, we recommend all future iterations of modeling crisis-severity consider severity as a multi-faceted construct. In doing so, practitioners should strive to create a parsimonious model. Inherently, humanitarian data are subject to high levels of uncertainty, and nonparsimonious models may further limit clear interpretation of severity within this context. Additionally, we recommend that future work consider longitudinal metrics of severity, as crises change within a given location over time. After further testing this model with additional crises, opportunities for converting model output to a dashboard or application for humanitarian actors should be explored. At the time of writing this manuscript, the current GCSI estimates were available in a large spreadsheet available at https://data.humdata.org/. They are now also available on the ACAPs website (https://www.acaps.org/methodology/severity) in an interactive dashboard and available to be accessed through an Application Programming Interface (API; https://api.acaps.org/). This interface provides a blueprint for merging robust statistical output with information needed by data users. With these recommendations in place, humanitarian actors can apply the humanitarian principle of impartiality when determining where need is the greatest and to best respond to crises.

## Data Availability

Data are publicly available.

https://data.humdata.org/dataset/inform-global-crisis-severity-index

## List of abbreviations

UNOCHA: United Nations Office for the Coordination of Humanitarian Affairs
UN: United Nations
GCSI: Global Crisis Severity Index
EFA: Exploratory Factor Analysis
CFA: Confirmatory Factor Analysis
CFI: Comparative Fit Index
TLI: Tucker-Lewis Index
RMSEA: Root Mean Square Error of Approximation
API: Application Programming Interface

## Declarations

### Ethics approval and consent to participate

Not applicable

### Consent for publication

Not applicable

### Availability of data and materials

The datasets generated and/or analysed during the current study are available in the *GCSI Database Beta Version – November 2019* repository, https://data.humdata.org/dataset/inform-global-crisis-severity-index

### Authors’ contributions

RG and VKL designed the research question, with assistance from AN. VKL and CB designed the analysis plan, and VKL conducted all analyses. VKL wrote the manuscript in consultation with RG, AN, CB, and LT. All authors provided critical feedback to help shape the analysis and manuscript.

## Acknowledgements

The following individuals provided value feedback on our methods and results: Mahlet Woldetsadik, Luca Vernaccini, Stefano Disperati, and Karmen Poljansek. We thank them for their insights, which helped to strengthen this analysis.

## CDC disclaimer

The findings and conclusions in this report are those of the authors and do not necessarily represent the official position of the Centers for Disease Control and Prevention.

