## Supplemental Materials for "Can Severity of a Humanitarian Crisis be Quantified? Assessment of the INFORM Severity Index"

**Appendix 1: The Global Crisis Severity Index (GCSI) Conceptual Framework and Estimation**

The GCSI is calculated as a weighted sum of three latent constructs: 1) the impact of the crisis, 2) the complexity of a crisis, and 3) the conditions of people as a result of the crisis. Each latent construct is referred to as a “pillar”. Each pillar is normalized on a scale from zero to five; however, pillar calculation is unique, as described below:

Impact of the Crisis: The impact of the crisis, which accounts for 20% of the final GCSI score, is the weighted sum of two composite indicators - The Human Impact (weighted at 0.7) and the Geographical Impact (weighted at 0.3).

The Human Impact is derived from averaging four indicators (People Affected, Displaced, Injured and Ill, and Fatalities), each of which is an average of sub-indictors. The People Affected indicator is the average of the absolute number of people affected and the percent of people affected relative to the total population. The People Displaced indicator is the average of the absolute number of people displaced and the percent of people displaced relative to the total population. The People Ill and Injured indicator is the average of the absolute number of people ill and the absolute number of people injured. Finally, the Fatalities indicator is a categorical version of the crude number of deaths that occurred.

The Geographical Impact is derived from averaging two indicators (Landmass Affected and People Living in an Affected Area), each of which is an average of sub-indictors. The Landmass Affected indicator is the average of the absolute value of land affected and the percent of landmass affected. The People Living in an Affected Area indicator is the average of the population size affected and the percent of the population living in the affected area.

All these sub-indicators are a categorical version (ranging from zero to five) of a raw, continuous variable.

Complexity of the Crisis: The complexity of the crisis, which accounts for 30% of the final GCSI score, is the geometric mean of two composite indicators – Society and Safety and Operating Environment.

The Society and Safety composite indicator is derived from averaging three indicators (Social Cohesion, Safety and Security, and Rule of Law), each of which is an average of sub-indictors. The Social Cohesion indicator is the average of the following three sub-indicators: Inequity, Ethnic Fractionalization, and Trust in Society. In turn, each of these sub-indicators are also averages of additional data points. Inequality is the average of Gender Inequality and the Income Gini Coefficient. Ethnic Fractionalization is the average of the size of excluded ethnic groups and a score of ethnic fractionalization within a country. Trust in Society is the average of a country’s empowerment score and democracy status. The Safety and Security sub-indicator is the average of conflict intensity and the total number of people killed in a crisis. The Rule of Law sub-indicator is the average of four indicators – corruption perception; rule of law (as measured by the Worldwide Government Indicators [WGI]), rule of law (as measured by the Bertelsmann Stiftung’s Transformation Index [BTI]), and freedom in the world.

The Operating Environment composite indicator is the average of two indictors, Humanitarian Access and Crisis Affected Groups. Humanitarian Access is the average of three sub-indicators (Physical and Security Constraints, Access of People in Need to Aid, and Access to Humanitarian Actors). Each of the sub-indicators is calculated by summing underlying data and ranking the total sum. The Physical and Security Constraints sub-indicator is a scaled sum of three variables – mines, insecurity, and environment; while the Access of People in Need to Aid is the sum of denial and obstructed access. Finally, Access to Humanitarian Actors sub-indicator is the sum of four variables- violence, impediments to entry, interference, and restricted movements.

Each of the data points used in the calculation for Complexity of Crisis are from publicly available sources. All indicators are scaled to range from 0 to 5 or have been transformed into categorical information using thresholds determined by the GCSI designers.

Conditions of the People as a Result of the Crisis: The conditions of the people as a result of the crisis accounts for 50% of the final GCSI score; it is the average of two sub-indicators – Current Humanitarian Conditions of the Total Population, and Current Humanitarian Conditions of the Population Affected.

Each of the sub-indicators ranks the percent of entire population or the population of affected people, respectively, into one of five levels: 1. those facing minimal humanitarian need, 2. those in stressed humanitarian conditions and needs, 3. those in moderate humanitarian conditions and needs; 4. those in severe humanitarian conditions and needs, and 5. those in extreme humanitarian conditions and needs.

For the total population, the Current Humanitarian Conditions of the Total Population sub-indicator reflects the sum of people in levels 3-5. This value is then categorized into a variable that ranges from zero to five. Categories were determined by INFORM designers.

The Current Humanitarian Conditions of the Population Affected sub-indictor, however, is calculated slightly differently. Here, the highest level is taken if the percent of the population affected at that level is greater than 5%. For example, if 66% of the population affected falls into level 2; 28% of population affected is in level 3, and 6% of the population affected is in level 4; the sub-indicator is given a value of 4.0

A schematic of the GCSI conceptual framework is presented in Appendix 1, Figure 1. Within the figure, each box is a data point, each oval a sub-indicator, and each circle a composite indicator.

Spearman rank correlation coefficients for all indicators are displayed in Appendix 1, Figure 2. High correlation values are observed within each pillar (denoted by boxes), as well as between the pillars.

**Appendix 1 Figure 1.** Diagram of Global Crisis Severity Index (GCSI) Model**.** Each box represents input data, which are aggregated in the methods described above into the oval shaped sub-indicators. Each circle represents a composite indicator.

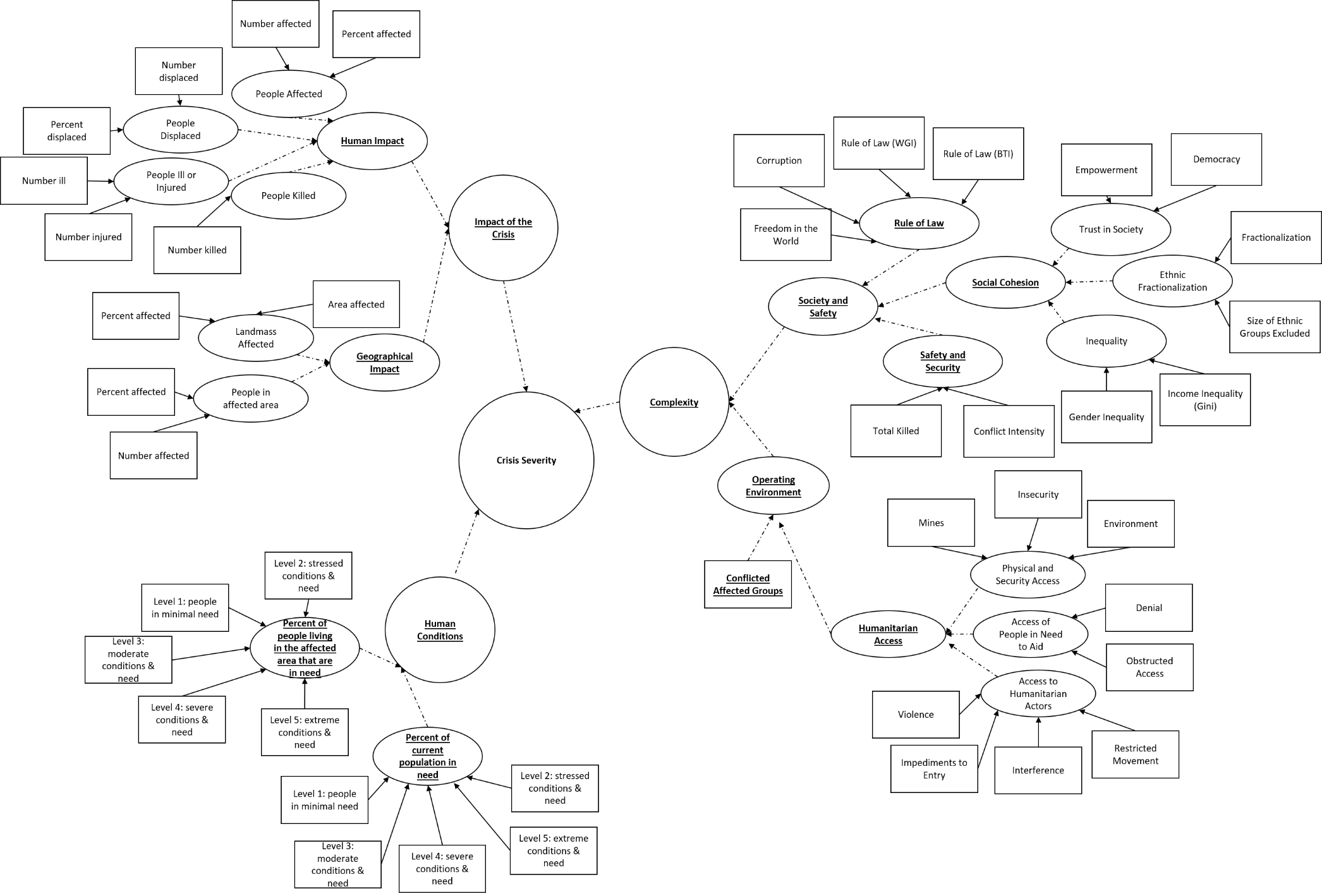

**Appendix 1 Figure 2.** Spearman rank correlation coefficients for all GCSI indicators. Indicators within the ‘Impact’ pillar are grouped with a grey box. Indicators within the ‘Complexity’ pillar are denoted with a dashed black line. Indicators in the ‘Human Conditions’ pillar are grouped with the solid black line.

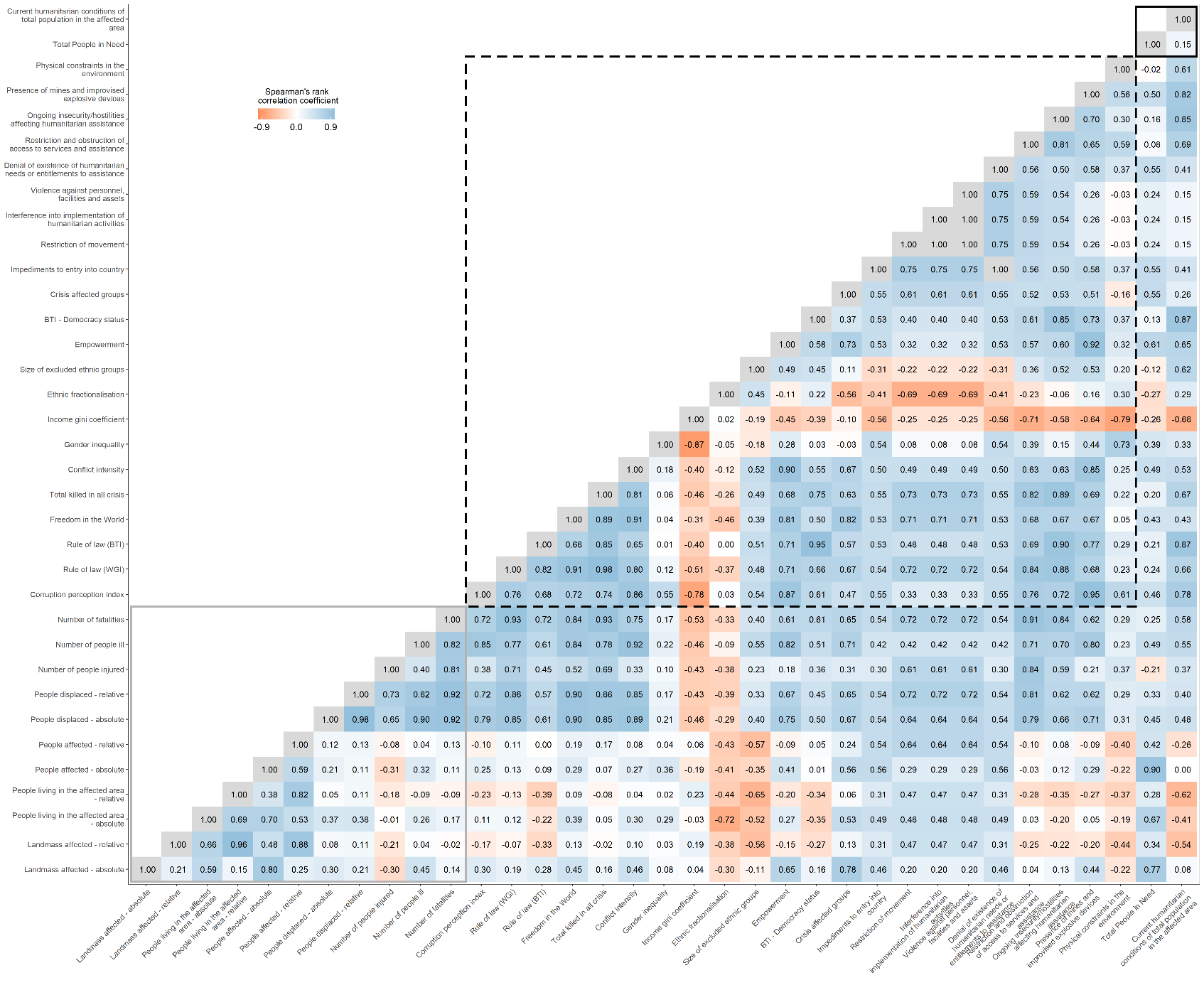

**Appendix 2: Factor loadings for 5-factor and 6-factor Exploratory Factor Analysis (EFA) solutions**

Factor loadings for indicators in the 5-factor and 6-factor EFA solutions are presented in Appendix Table 2.1. Cross-loadings were common, suggesting that these indicators provide negligible contributions to the factors. For example, in the 5-factor solution, of the 7 indicators that were grouped into *factor 1*, four had substantial cross-loadings onto other factors, while of the 10 indicators contributing to *factor 5*, 6 also had cross-loadings with other factors within the solution. This pattern was also evident in the 6-factor solution. Here, in *factor 2*, one-third of indicators were cross-loaded, while for factor 4, three-fifths of indicators showed high cross-loadings.

| **Appendix Table 2.1.** Global Crisis Severity Index (GCSI) Indicators factor loadings for each Exploratory Factor Analysis (EFA) factor solution. Factor loadings are only presented if greater than the absolute value of 0.3 (*GCSI dataset, 2019, N=172*). | | | | | | | | | | | |
| --- | --- | --- | --- | --- | --- | --- | --- | --- | --- | --- | --- |
| **Indicators** | **Five-Factor Solution** | | | | | **Six-Factor Solution** | | | | | |
|  | *Factor 1* | *Factor 2* | *Factor 3* | *Factor 4* | *Factor 5* | *Factor 1* | *Factor 2* | *Factor 3* | *Factor 4* | *Factor 5* | *Factor 6* |
| Landmass affected - absolute | 0.59 |  |  |  |  |  |  |  | 0.53 |  |  |
| Landmass affected - relative | 0.32 |  | 0.57 |  |  |  | 0.86 |  |  |  |  |
| People living in the affected area - absolute | 0.97 |  |  |  |  |  |  |  | 0.80 |  |  |
| People living in the affected area - relative | 0.31 |  | 0.66 |  |  |  | 0.93 |  |  |  |  |
| People affected - absolute |  |  | 0.60 |  |  |  |  |  |  | 0.92 |  |
| People affected - relative |  |  | 0.98 |  |  |  | 0.40 |  | -0.32 | 0.72 |  |
| People displaced - absolute |  |  |  | 0.35 | -0.38 |  |  |  |  | 0.48 | -0.32 |
| People displaced - relative |  |  | 0.48 |  | -0.37 |  |  |  |  | 0.31 | -0.37 |
| Corruption Perception |  | 0.56 |  |  | 0.44 | 0.56 |  |  |  |  | 0.41 |
| Rule of Law (WGI) |  | 0.45 |  | 0.40 | 0.44 | 0.45 |  | 0.42 |  |  | 0.40 |
| Rule of Law (BTI) |  | 0.96 |  |  |  | 0.95 |  |  |  |  |  |
| Freedom in the world |  | 0.88 |  |  |  | 0.88 |  |  |  |  |  |
| Total killed in all crisis |  |  |  | 0.63 |  |  |  | 0.61 |  |  |  |
| Conflict intensity | 0.30 |  | -0.30 | 0.59 |  |  |  | 0.53 | 0.37 |  |  |
| Gender inequality |  |  |  |  | 0.54 |  |  |  |  |  | 0.60 |
| Income gini coefficient |  |  |  |  | 0.41 |  |  |  |  |  | 0.36 |
| Ethnic fractionalisation |  |  |  |  | 0.33 |  |  |  |  |  | 0.41 |
| Size of excluded ethnic groups |  | 0.38 |  |  |  | 0.38 |  | -0.35 |  |  |  |
| Empowerment |  | 0.57 |  |  |  | 0.56 |  |  |  |  |  |
| BTI - Democracy Status |  | 0.95 |  |  |  | 0.94 |  |  |  |  |  |
| Crisis affected groups |  |  |  | 0.48 |  |  |  | 0.44 |  |  |  |
| Impediments to entry into country (bureaucratic and administrative) |  | 0.43 |  | 0.32 | -0.37 | 0.43 |  | 0.32 |  |  | -0.39 |
| Restriction of movement (impediments to freedom of movement and/or administrative restrictions) |  |  |  | 0.93 |  |  |  | 0.96 |  |  |  |
| Interference into implementation of humanitarian activities |  |  |  | 0.68 |  |  |  | 0.72 |  |  |  |
| Violence against personnel, facilities and assets |  |  |  | 0.55 |  |  |  | 0.53 |  |  |  |
| Denial of existence of humanitarian needs or entitlements to assistance |  | 0.31 |  |  | -0.43 | 0.30 |  | 0.33 |  |  | -0.47 |
| Restriction and obstruction of access to services and assistance |  |  |  | 0.78 |  |  |  | 0.77 |  |  |  |
| Ongoing insecurity/hostilities affecting humanitarian assistance |  |  |  | 0.83 |  |  |  | 0.82 |  |  |  |
| Presence of mines and improvised explosive devices |  |  |  | 0.58 |  |  |  | 0.54 |  |  |  |
| Physical constraints in the environment (obstacles related to terrain, climate, lack of infrastructure, etc.) |  |  |  | 0.31 | 0.31 |  |  | 0.35 |  |  |  |
| Total People in Need | 0.40 |  | 0.48 |  |  |  |  |  |  | 0.74 |  |
| Current humanitarian conditions of total population in the affected area | -0.31 |  | 0.36 |  |  |  |  |  | -0.35 | 0.47 |  |

**Appendix 3: Correlation Matrix for First-Order Confirmatory Factor Analysis (CFA)**

| **Appendix Table 3.1**. Residual correlations for all indicators included in the Confirmatory Factor Analysis (CFA) with three latent factors. Indicators modeled on the same factor are depicted with a grey box. | | | | | | | | | | | | | | | | | | | | | | |
| --- | --- | --- | --- | --- | --- | --- | --- | --- | --- | --- | --- | --- | --- | --- | --- | --- | --- | --- | --- | --- | --- | --- |
|  | land_rel | pop_rel | ppl_rel | dis_rel | corrupt | law_BTI | free | gender | emp | bti | environ | killed | CI | gini | CAG_Indicator | restr | interfer | violence | ob.access | insecure | mines | dis_ab |
| land_rel | 0.00 |  |  |  |  |  |  |  |  |  |  |  |  |  |  |  |  |  |  |  |  |  |
| pop_rel | 0.01 | 0.00 |  |  |  |  |  |  |  |  |  |  |  |  |  |  |  |  |  |  |  |  |
| ppl_rel | -0.03 | -0.01 | 0.00 |  |  |  |  |  |  |  |  |  |  |  |  |  |  |  |  |  |  |  |
| dis_rel | -0.08 | -0.03 | 0.21 | 0.00 |  |  |  |  |  |  |  |  |  |  |  |  |  |  |  |  |  |  |
| corrupt | 0.06 | 0.02 | 0.19 | 0.18 | 0.00 |  |  |  |  |  |  |  |  |  |  |  |  |  |  |  |  |  |
| law_BTI | 0.00 | -0.02 | 0.14 | 0.20 | -0.01 | 0.00 |  |  |  |  |  |  |  |  |  |  |  |  |  |  |  |  |
| free | 0.02 | -0.01 | 0.14 | 0.31 | 0.02 | 0.00 | 0.00 |  |  |  |  |  |  |  |  |  |  |  |  |  |  |  |
| gender | 0.01 | -0.03 | 0.15 | -0.07 | 0.15 | -0.07 | 0.01 | 0.00 |  |  |  |  |  |  |  |  |  |  |  |  |  |  |
| emp | 0.00 | -0.11 | 0.00 | 0.13 | -0.07 | -0.04 | 0.04 | -0.10 | 0.00 |  |  |  |  |  |  |  |  |  |  |  |  |  |
| bti | -0.05 | -0.09 | 0.08 | 0.16 | -0.07 | 0.01 | -0.02 | -0.06 | 0.12 | 0.00 |  |  |  |  |  |  |  |  |  |  |  |  |
| environ | 0.07 | 0.01 | 0.08 | 0.05 | 0.14 | -0.01 | -0.03 | 0.14 | -0.09 | -0.05 | 0.00 |  |  |  |  |  |  |  |  |  |  |  |
| killed | 0.01 | -0.08 | -0.01 | 0.12 | 0.08 | -0.01 | -0.10 | -0.05 | 0.05 | -0.03 | 0.15 | 0.00 |  |  |  |  |  |  |  |  |  |  |
| CI | -0.12 | -0.19 | -0.15 | 0.12 | 0.16 | 0.02 | 0.12 | 0.14 | 0.13 | 0.03 | 0.07 | 0.09 | 0.00 |  |  |  |  |  |  |  |  |  |
| gini | 0.14 | 0.23 | 0.07 | -0.11 | 0.02 | -0.07 | -0.04 | 0.11 | -0.38 | -0.18 | 0.04 | 0.10 | 0.03 | 0.00 |  |  |  |  |  |  |  |  |
| CAG_Indicator | 0.23 | 0.17 | 0.30 | 0.21 | 0.10 | -0.04 | 0.06 | 0.22 | -0.01 | -0.05 | 0.22 | 0.04 | 0.12 | 0.07 | 0.00 |  |  |  |  |  |  |  |
| restr | 0.03 | -0.03 | 0.15 | 0.28 | 0.16 | -0.06 | 0.01 | 0.05 | 0.06 | -0.07 | 0.17 | -0.03 | -0.05 | -0.01 | -0.01 | 0.00 |  |  |  |  |  |  |
| interfer | 0.10 | 0.07 | 0.14 | 0.38 | 0.13 | 0.04 | 0.11 | -0.11 | 0.14 | 0.00 | 0.15 | -0.08 | -0.07 | 0.02 | -0.10 | 0.04 | 0.00 |  |  |  |  |  |
| violence | 0.02 | 0.02 | 0.13 | 0.13 | 0.11 | -0.04 | 0.00 | 0.25 | 0.12 | 0.06 | 0.06 | 0.08 | -0.01 | 0.02 | 0.02 | 0.02 | -0.04 | 0.00 |  |  |  |  |
| ob.access | 0.05 | -0.03 | 0.12 | 0.33 | 0.00 | -0.12 | -0.13 | 0.00 | 0.02 | -0.12 | 0.17 | 0.08 | -0.01 | -0.03 | 0.01 | 0.00 | 0.00 | -0.07 | 0.00 |  |  |  |
| insecure | -0.02 | -0.07 | 0.07 | 0.17 | 0.15 | 0.00 | 0.00 | 0.16 | -0.04 | -0.04 | 0.30 | -0.03 | 0.01 | 0.09 | -0.02 | 0.02 | -0.01 | 0.03 | 0.01 | 0.00 |  |  |
| mines | -0.06 | -0.10 | -0.02 | 0.24 | 0.23 | 0.03 | 0.13 | 0.05 | 0.11 | 0.02 | 0.08 | 0.09 | 0.18 | -0.03 | 0.01 | -0.02 | -0.03 | 0.01 | -0.05 | -0.03 | 0.00 |  |
| dis_ab | 0.08 | 0.15 | 0.30 | 0.72 | 0.02 | -0.02 | 0.07 | -0.14 | 0.07 | -0.01 | -0.12 | 0.03 | 0.13 | 0.01 | 0.12 | -0.05 | 0.04 | -0.06 | 0.09 | -0.06 | 0.05 | 0.00 |

Appendix Table 3.1 shows the correlation matrix for all indicators in the CFA with 3 latent constructs. In this table, the indicators are shaded to represent the latent constructs that they represent within the model. We applied model building by removing indicators if they had residual correlation greater than 0.10 with indicators across different latent constructs. As evident in the table, many of the indicators showed strong correlation patterns with multiple indicators.

**Appendix 4: R code for primary analyses**

###Libraries

library(psych)

library(GPArotation)

library(lavaan)

####---- General EFA: Multifactor ----####

##Remove ill, injury, and fatality variables (Due to lots of missing information)

indicators2 <-subset(indicators, select = -c(injur, ill, fatal))

#All possible variable/factor relationships

#Identify patterns in the data

#Review of eigenvalues

bfi_EFA_cor <-cor(indicators2, use="pairwise.complete.obs")

eigenvals <-eigen(bfi_EFA_cor) #>1 are meaningful factors

scree(bfi_EFA_cor, factors=FALSE) #Scree suggests 5 factors; should try 3

M3 <-fa(indicators2, nfactors=3, fm="ml", missing=TRUE, impute="median")

M3

print(M3$loadings, cutoff=0.3)

M4 <-fa(indicators2, nfactors=4, fm="ml", missing=TRUE, impute="median")

M4

print(M4$loadings, cutoff=0.3)

M5 <-fa(indicators2, nfactors=5, fm="ml", missing=TRUE, impute="median")

M5

print(M5$loadings, cutoff=0.3)

M6 <-fa(indicators2, nfactors=6, fm="ml", missing=TRUE, impute="median")

M6

print(M6$loadings, cutoff=0.3)

####---- CFAs (from EFAs) ----####

###3 Factor CFA

CFA_3 <-'

F2 =~ land_rel + pop_rel + ppl_rel + dis_rel

F1 =~ corrupt + law_WGI + law_BTI + free + gender + emp + bti + environ

F3 =~ killed + CI + gini + CAG_Indicator + restr + interfer + violence + ob.access + insecure + mines

'

fit_3 <-cfa(CFA_3, data=gcsi_nov2019, missing="ML")

summary(fit_3, standardize=TRUE)

fitMeasures(fit_3)

parameterEstimates(fit_3, standardized=TRUE)

###4 Factor CFA

CFA_4 <-'

F2 =~ land_rel + pop_rel + ppl_rel + dis_rel

F1 =~ corrupt + law_BTI + free + emp + bti

F3 =~ killed + CI + CAG_Indicator + restr + interfer + violence + ob.access + insecure + mines

F4 =~ gender + environ

'

fit_4 <-cfa(CFA_4, data=gcsi_nov2019, std.ov=TRUE, missing="ML")

summary(fit_4, standardize=TRUE)

fitMeasures(fit_4)

parameterEstimates(fit_4, standardized=TRUE)

###5 Factor CFA

CFA_5 <-'

F2 =~ corrupt + law_BTI + free + eth_exclude + emp + bti

F1 =~ land_ab + pop_ab

F3 =~ ppl_ab + ppl_rel

F4 =~ killed + CI + CAG_Indicator + restr + interfer + violence + ob.access + insecure + mines

F5 =~ gender + gini + ef

'

fit_5 <-cfa(CFA_5, data=gcsi_nov2019, missing="ML") #Error: No solution found

###6 Factor CFA

CFA_6 <-'

F2 =~ land_rel + pop_rel

F1 =~ law_BTI + free + emp + bti

F3 =~ killed + CAG_Indicator + restr + interfer + violence + ob.access + insecure + mines

F4 =~ land_ab + pop_ab

F5 =~ ppl_ab + ppl_need

F6 =~ gender + gini + ef

'

fit_6 <-cfa(CFA_6, data=gcsi_nov2019) #Error: No solution found

\#####

####---- CFA with 3 LVs ----####

CFA_3B <-'

F2 =~ land_rel + pop_rel + ppl_rel

F1 =~ law_BTI + free + gender + emp + bti

F3 =~ killed + restr + ob.access

gender ~~ emp

'

fit_3B <-cfa(CFA_3B, data=gcsi_nov2019, std.ov=TRUE, missing="ML")

summary(fit_3B, standardize=TRUE)

fitMeasures(fit_3B)

resid(fit_3B, type="cor")

##Second-order LV for severity

CFA_3C <-'

GCSI =~ F1 + F2 + F3

F2 =~ land_rel + pop_rel + ppl_rel

F1 =~ law_BTI + free + gender + emp + bti

F3 =~ killed + restr + ob.access

gender ~~ emp

'

fit_3C <-cfa(CFA_3C, data=gcsi_nov2019, std.ov=TRUE, missing="ML")

summary(fit_3C, standardize=TRUE)

fitMeasures(fit_3C)

resid(fit_3C, type="cor")

####---- CFA with 4 LVs ----####

CFA_4B <-'

F2 =~ land_rel + pop_rel + ppl_rel

F1 =~ law_BTI + free + bti

F3 =~ killed + restr + ob.access + insecure

F4 =~ gender + environ

killed ~~ ob.access

'

fit_4D <-cfa(CFA_4B, data=gcsi_nov2019, std.ov=TRUE, missing="ML")

summary(fit_4B, standardize=TRUE)

fitMeasures(fit_4B)

resid(fit_4B, type="cor")

##Add second-order LV for severity

CFA_4C <-'

GCSI =~ F1 + F2 + F3

F2 =~ land_rel + pop_rel + ppl_rel

F1 =~ law_BTI + free + bti

F3 =~ killed + restr + ob.access + insecure

F4 =~ gender + environ

killed ~~ ob.access

'

fit_4E <-cfa(CFA_4C, data=gcsi_nov2019, std.ov=TRUE, missing="ML")

summary(fit_4C, standardize=TRUE)

fitMeasures(fit_4C)

resid(fit_4E, type="cor")

**Appendix 5: Inclusion of “Conditions of People Affected by the Crisis”**

Calculating the number of people in need of humanitarian assistance is standard metric of providing humanitarian assistance. The most common data sources include humanitarian needs assessment, an Integrated Phase Classification (IPC) assessment or *cadre harmonise*, displacement data collected in the International Organization for Migration’s (IOM) Displacement Tracking Matrix (DTM) tool, and information provided from United National High Commissioner for Refugees (UNHCR). In addition, if available, Humanitarian Response Plans (HRPs) and Humanitarian Needs Overview (HNOs), which are reports that compile multiple data sources on humanitarian crises, are usually prioritized as a source. However, despite their programmatic importance, indicators reflecting people in need, but were removed from our model building process. Because such calculations are standard in distribution of humanitarian aid, we re-ran the second-order Confirmatory Factor Analysis (CFA) model and included the two ‘condition of the people’ indicators^[1]^ as standalone independent variables (rather than latent constructs, since the two indicators are not correlated). Specifically, we ran the following models CFAs:

- Model 1: Second-order CFA + People in Need + People Affected
- Model 2: Second-order CFA + People in Need
- Model 3: Second-order CFA + People Affected

We compare the model diagnostics of the above model to the second-order CFA in the main text (Appendix Table 5.1). Additionally, we examined descriptive statistics for the severity score calculated by each model; we also evaluated the Pearson correlation coefficient and intraclass correlation coefficient (ICC) of each model to that of the second-order CFA in the main text (Appendix Table 5.2). Specifically, we examined an ICC for agreement with two-way mixed affects and fixed raters. The correlations provide insight into the reliability and agreement of the results.

| **Appendix Table 5.1.** Fit statistics and Factor ladings for each model. Fit statistics include Chi-squared goodness of fit test statistic, Comparative Fit Index (CFI), Tucker-Lewis Index (TLI), and Root Mean Square Error of Approximation (RMSEA) Index (*GCSI dataset, 2019, N=172*). | | | | | | | | | |
| --- | --- | --- | --- | --- | --- | --- | --- | --- | --- |
| **Models** | **Model Fit** | | | | **Factor Loadings** | | | | |
|  | **Chi-squared goodness of fit (degrees of freedom)** | **CFI** | **TLI** | **RMSEA** | **Societal Governance** | **Humanitarian access/safety** | **Impact** | **People in need** | **People affected** |
| Model 1 | 191 (61) | 0.894 | 0.865 | 0.094 | 0.40 | 0.51 | 0.44 | 0.84 | 0.48 |
| Model 2 | 159 (50) | 0.904 | 0.879 | 0.093 | 0.40 | 0.53 | 0.48 | 0.78 | - |
| Model 3 | 137 (50) | 0.924 | 0.900 | 0.101 | 0.77 | 0.55 | 0.20 | - | 0.41 |
| Model in main text | 107 (40) | 0.940 | 0.917 | 0.099 | 0.73 | 0.59 | 0.21 | - | - |

| **Appendix Table 5.2.** Descriptive statistics for latent severity crisis scores in each model: Median, mean, Pearson correlation coefficient, and intraclass correlation coefficient (ICC), with 95% confidence interval (CI) (*GCSI dataset, 2019, N=172*). | | | | |
| --- | --- | --- | --- | --- |
| **Models** | **Median** | **Mean** | **Pearson correlation coefficient** | **ICC (95% CI)** |
| Model 1 | 0.45 | 0.47 | 0.63 | 0.63 (0.53-0.71) |
| Model 2 | 0.44 | 0.05 | 0.67 | 0.67 (0.58-0.75) |
| Model 3 | 0.54 | 0.54 | 0.98 | 0.98 (0.97-0.98) |
| Model in main text | 0.53 | 0.54 | - | - |

Of the models, model 3 had the best fit and was comparable to the second-order CFA (Appendix Table 5.1). In addition, the factor loadings were comparable between these two models. Further evidence of comparability is provided by the descriptive statistics (Appendix Table 5.2); the severity scores from model 3 exhibited strong correlation and agreement with the final model of the main text (Pearson’s correlation coefficient=0.98; ICC=0.98 [95% CI 0.97-0.99]). Collectively, these results suggest that including the indicator for People Affected will generate comparable results to a model that excludes this information.

**Appendix 6: Indicator reliability**

Analysts that collect the Global Crisis Severity Index (GCSI) data rank indicators within each crisis for reliability. Reliability is qualitatively scored as “high”, “medium”, or “low”. The data underlying the indicators in the “Conditions of People Affected by the Crisis” pillar have generally lower reliability (greater than 20% of crises are scored low for these indicators) relative to other indicators (Appendix Table 6.1). In addition, indicators related to counting deaths, such as ‘number of fatalities’ and ‘total number of killed in all crisis’ are also subject to lower reliability.

| **Appendix Table 6.1**. Distribution of indicator reliability, as scored by GCSI analysts. | | | | |
| --- | --- | --- | --- | --- |
|  | High | Medium | Low | N |
| Landmass affected - absolute* | 0.20 | 0.74 | 0.06 | 162 |
| People living in the affected area - absolute* | 0.09 | 0.80 | 0.11 | 161 |
| People affected - absolute* | 0.04 | 0.78 | 0.18 | 155 |
| People displaced - absolute** | 0.10 | 0.71 | 0.19 | 140 |
| Number of people ill | 0.08 | 0.85 | 0.07 | 87 |
| Number of people injured | 0.05 | 0.90 | 0.05 | 78 |
| Number of fatalities | 0.08 | 0.69 | 0.23 | 131 |
| Level 1: None/Minor humanitarian conditions | 0.02 | 0.76 | 0.22 | 145 |
| Level 2: Stressed humanitarian conditions | 0.02 | 0.77 | 0.21 | 143 |
| Level 3: Moderate humanitarian conditions | 0.01 | 0.73 | 0.26 | 143 |
| Level 4: Severe humanitarian conditions | 0.02 | 0.73 | 0.25 | 141 |
| Level 5: Extreme humanitarian conditions | 0.02 | 0.77 | 0.21 | 141 |
| Total killed in all crisis* | 0.08 | 0.62 | 0.30 | 145 |
| Crisis affected groups** | 0.27 | 0.71 | 0.02 | 150 |
| Impediments to entry into country (bureaucratic and administrative)** | 0.51 | 0.48 | 0.01 | 161 |
| Restriction of movement (impediments to freedom of movement and/or administrative restrictions)* | 0.45 | 0.48 | 0.06 | 163 |
| Interference into implementation of humanitarian activities** | 0.43 | 0.55 | 0.02 | 163 |
| Violence against personnel, facilities and assets** | 0.46 | 0.49 | 0.05 | 165 |
| Denial of existence of humanitarian needs or entitlements to assistance** | 0.45 | 0.51 | 0.04 | 159 |
| Restriction and obstruction of access to services and assistance* | 0.48 | 0.49 | 0.04 | 162 |
| Ongoing insecurity/hostilities affecting humanitarian assistance** | 0.48 | 0.49 | 0.02 | 164 |
| Presence of mines and improvised explosive devices ** | 0.44 | 0.54 | 0.03 | 157 |
| Physical constraints in the environment (obstacles related to terrain, climate, lack of infrastructure, etc.)** | 0.46 | 0.54 | 0.00 | 162 |
| **Indicator retained in final model* | | | | |
| ***Indicator removed from final model* | | | | |

We compared the reliability of the indicators retained in the final model to the reliability of indicators that were removed. To do so, for each indicator, we added the total number of crises that were scored as low, medium, or high reliability. We then constructed three logistic regression models to assess the association between whether the indicator was retained and each reliability level. We found that retained indicators had a marginally lower likelihood of having high reliability scores (Appendix Table 6.2), but showed no difference between medium and low reliability scores.

| **Appendix Table 6.2**. Logistic regression models for retained indicators (outcome) and reliability score. Beta estimates are shown for retained indicators, given the number of crises with each reliability score | | | |
| --- | --- | --- | --- |
| Reliability Score | Beta | Standard Error | p-value |
| Low | 0.12 | 0.07 | 0.10 |
| Medium | 0.06 | 0.04 | 0.11 |
| High | -0.04 | 0.02 | 0.08 |

**Appendix 7: Sensitivity analyses for derived latent severity scores**

We compared the latent severity score derived from our models to the original GCSI score by first plotting the scores relative to each other (Appendix 7 Figure 1A), and then by comparing the density distribution of each score (Appendix 7 Figure 1B and Appendix 7 Figure 1C).

**Appendix 7 Figure 1.** Comparison of latent crisis severity scores with original GCSI scores. Panel A shows a scatter plot of the scores, color coded by crisis type. Panels B and C are the density plot of the score distribution for each method of derivation, with the median score marked by a dashed line.

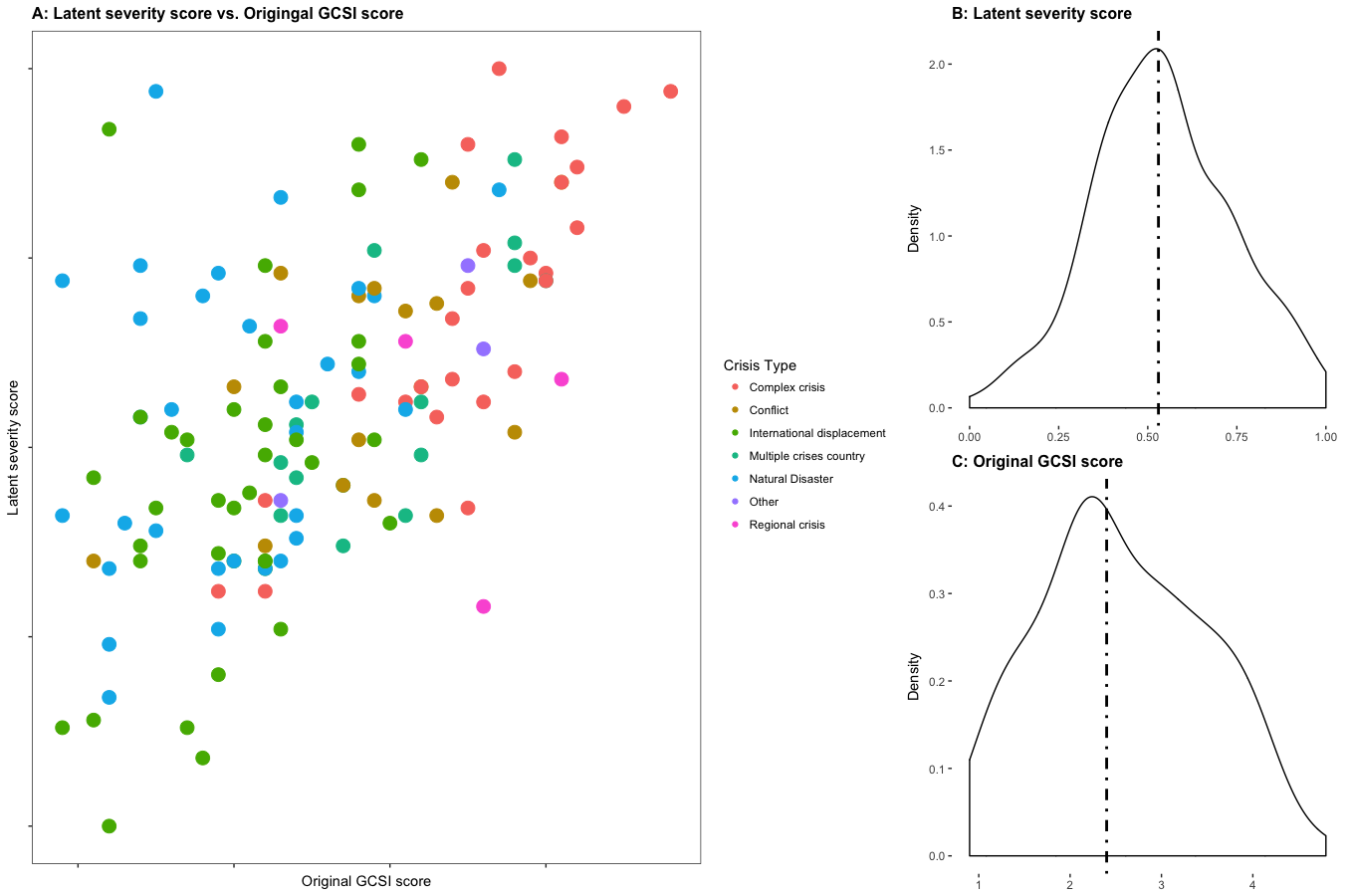

The scatter plot in Appendix 7 Figure 1A shows how well the two derivations of scores are correlated. If highly correlated, which implies they are equivalent metrics, we would expect the values to group along the diagonal (from the bottom left to the top right) of the figure. However, Appendix 7 Figure 1A shows no apparent pattern for most crises, with better correlation for complex crises. This pattern is not surprising when the distributions of severity scores are compared between the two approaches. Appendix 7 Figure 1B shows density plot of the latent severity scores, while Appendix 7 Figure 1C shows the density plot of the original GCSI scores. The latent scores have a near normal distribution, while the original GCSI scores have distribution that skews to the right. In other words, the original GCSI tends to score crises at lower values than the Confirmatory Factor Analysis (CFA) models. Conceptually, we would expect that crisis severity scores to present a normal distribution because the scores are a relative representation of crises. Thus, it is likely that the score derived from the CFA models is a closer representation of severity than those calculated from the GCSI methodology.

**Appendix 8: The role of state fragility in the final model**

As shown in the main text, indicators reflecting state fragility are a main driver of crisis severity. To test whether our model discriminates between crisis severity and state fragility, we assessed how the model scored countries with fragile states, but without crisis in our simulation. This sensitivity analysis had three main steps:

1. Incorporate additional data: We first randomly selected 10 GCSI crises with “Complexity” scores (this is the GCSI pillar that contains fragility-related indicators) that were greater than 2.6 (the median value for that pillar). These observations accounted for approximately 6% (10/182) of the new sample. We then recoded all non-fragility related indicators to 1 or 0, following classification schemes used by the GCSI creators. We chose not to set all values to zero in order to allow for slight variation, which is useful for the CFA. It should be noted that when we sampled more than 10 crises, the limited variance in the non-fragility related indicators dramatically impacted the model convergence.
2. Rerun the final CFA model: After merging the additional data with the entire dataset (N=182), we reran the final CFA model presented in the main text and generated scores for latent severity.
3. Compare scores: We compared the estimated severity scores for the 10 added observations to the remainder of the data. In theory, these scores should be lower.

Overall, we found the 10 simulated “non-emergencies” had much lower mean severity scores relative to the actual emergencies in the dataset (0.06 and 0.43, respectively). This suggests that the model structure does indeed allow for disaggregation between “fragility” and “severity”.

**Appendix 9: Missing data exploration**

When data are missing at random, inappropriate adjust for misvalues may bias a model’s results. In our main paper, we replaced missing values for the Exploratory Factor Analysis (EFA) with the median from the distribution of that variable and for the Confirmatory Factor Analysis (CFA), we used maximum likelihood (ML) to address the missing values. Here, we further explore the implication of these decisions by first rerunning the EFA model with case deletion and then rerunning the CFA model with multiple imputation (MI). Results from each approach is then compared to the results presented in the main paper.

EFA missing data exploration: We created an EFA model to guide decision making for the CFA model; hence, we sought to include as much information as possible in the main paper and imputed missing observations to the median value of the given indicator distribution. To test whether this decision had a large impact on the EFA results, we rerun the 3-factor solution EFA with case deletion. We chose to run the 3-factor model, rather than all six models (as done in the main paper analyses), since this was the model that we ultimately used to inform the CFA. Factor loadings for the 3-factor EFA with case deletion relative to the 3-factor EFA with imputed missing data are presented in Appendix 9 Figure 1.

**Appendix 9 Figure 1.** Factor loadings for 3-factor EFA model with case deletion (in red) and with median imputed missing values (in orange).

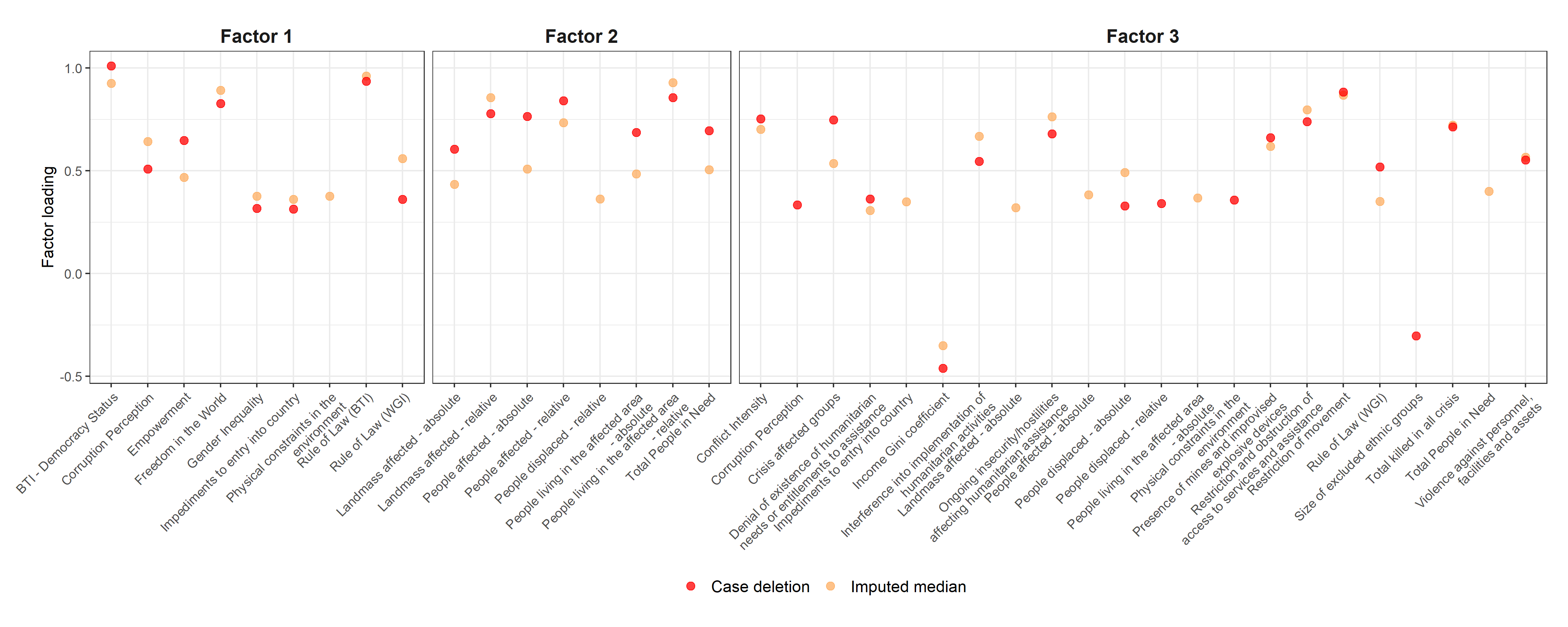

The factor loadings for the 3-factor EFA with case deletion (in red) and the 3-factor EFA with imputed missing data (in orange) are strikingly similar for factor 1 and factor 2. The model with imputed data had two indicators here that were not included in the model with case deletion: *Physical constraints in the environment* mapped on to factor 1 and the *relative number of displace people* mapped onto to factor 2. As both indicators were later deemed uninformative during CFA, the discrepancy between the factor loading for each missing data approach is likely not meaningful.

In contrast to the first two factors, factor 3 showed some differences in which variables may be important. In factor 3, three indicators were included in the case deletion model, but not the median imputation model: *Corruption perception*, *Physical constraints in the environment*, and *Size of excluded ethnic groups*. Nevertheless, we ultimately would not have included these three indicators in the final CFA model because *physical constraints in the environment* and the *size of excluded ethnic groups* had residual correlation greater than 0.1 with indicators on other factors (when the CFA model included these indicators) and *Corruption perception* had EFA cross-loadings on factor 1.

CFA missing data exploration: Our main paper analyses using maximum likelihood (ML) to fit the CFA models, which uses each case’s available data to compute ML estimates. This approach is a standard application when data are either missing completely at random or missing at random. To assess whether this approach was adequate to address the missing data in our sample, we compared it to results from a CFA model run with multiple imputation (MI). Appendix 9 Figure 2 shows model fit statistics (Comparative Fit Index (CFI), Tucker-Lewis Index (TLI), and Root Mean Square Error of Approximation (RMSEA)) from each missing data approach in Panel A and factor loadings from each model in Panel B.

**Appendix 9 Figure 2.** CFI, RMSEA with 95% confidence intervals, and TLI for the CFA models run with ML (in pink) and ML (in purple) are in Panel A. Standardized factor loadings for the MI model and standardized factor loadings with 95% confidence intervals for the ML model are in Panel B.

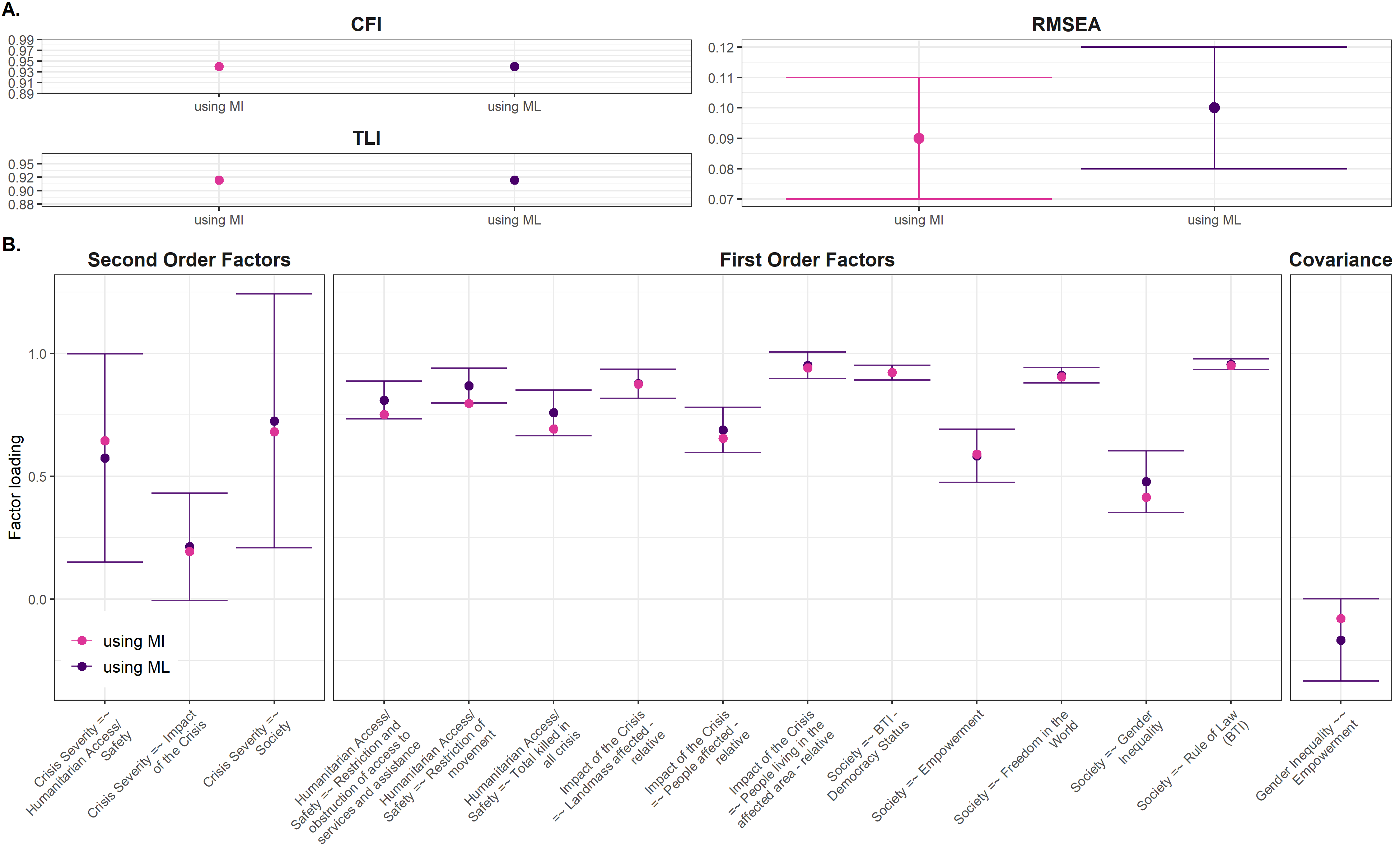

The MI (denoted in pink in the figure) approach generated a model with comparable fit to the ML model (denoted in purple in the figure). The MI model, nevertheless, had a slightly lower chi-squared value (95 vs 107, respectively). Standardized factor loadings from the MI model were within the 95% confidence intervals of the CFA run with ML.

Conclusion: Overall, these analyses suggest that missing indicators were unlikely to be missing at random and our results unlikely to be biased. Thus, the approaches we used to address missing observations in the main paper did not impact the accuracy of our findings.

**Appendix 10: Further assessment of model fit**

To determine whether the final Confirmatory Factor Analysis (CFA) model was overfit to the data, we created 1000 samples of 150 observations (without replacement) and re-ran the final CFA. We chose not to create samples of 172 observations (the full size of the dataset) given the potentially limited variance that would occur in some of the samples and inhibit model convergence. Our decision to draw samples of 150 observations was informed by Muthen and Muthen’s sample size guidance^[2]^. Appendix Figure 10.1 shows the distribution of model fit statistics (chi-square goodness-of-fit statistic, Comparative Fit Index (CFI), Tucker-Lewis Index (TLI), and Root Mean Square Error of Approximation (RMSEA)) from the 1000 samples overlaid with red points reflecting the fit statistics from the main paper findings; and Appendix Figure 10.2 shows the distribution of factor loadings from the 1000 samples and the main paper findings denoted as red points. Two samples were suppressed from Appendix Figure 10.2 given outlier factor loadings greater than 3.0 for Society ~ Crisis Severity.

**Appendix 10 Figure 1.** Distribution of CFI, chis-squared goodness of fit statistic (CHISQ), RMSEA, and TLI for the 1000 CFA models and main paper CFA factor loadings in red points.

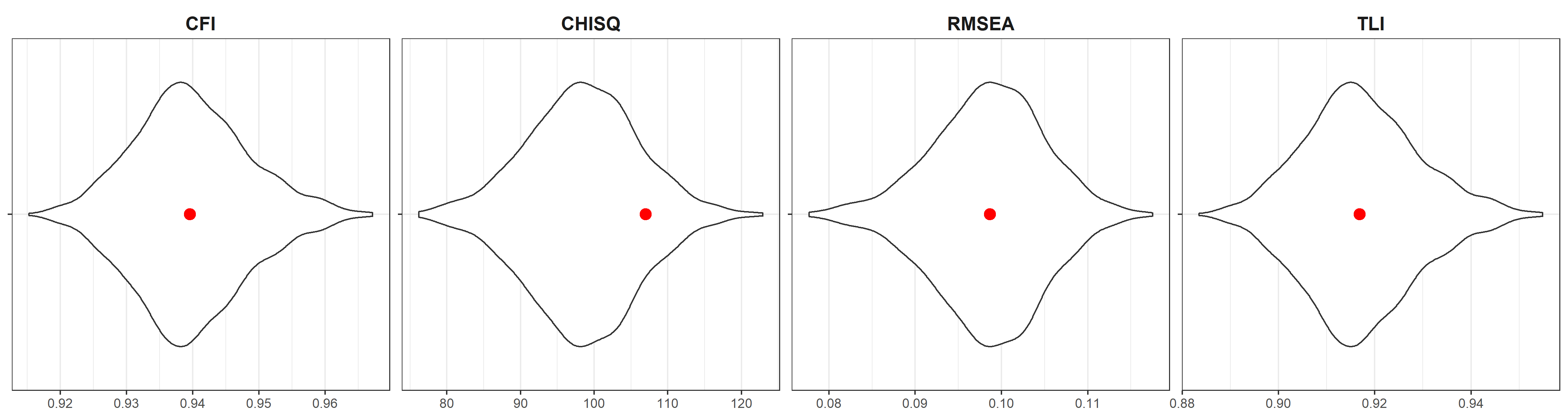

**Appendix 10 Figure 2.** Distribution of factor loadings estimated for the 1000 CFA models and main paper CFA factor loadings in red points.

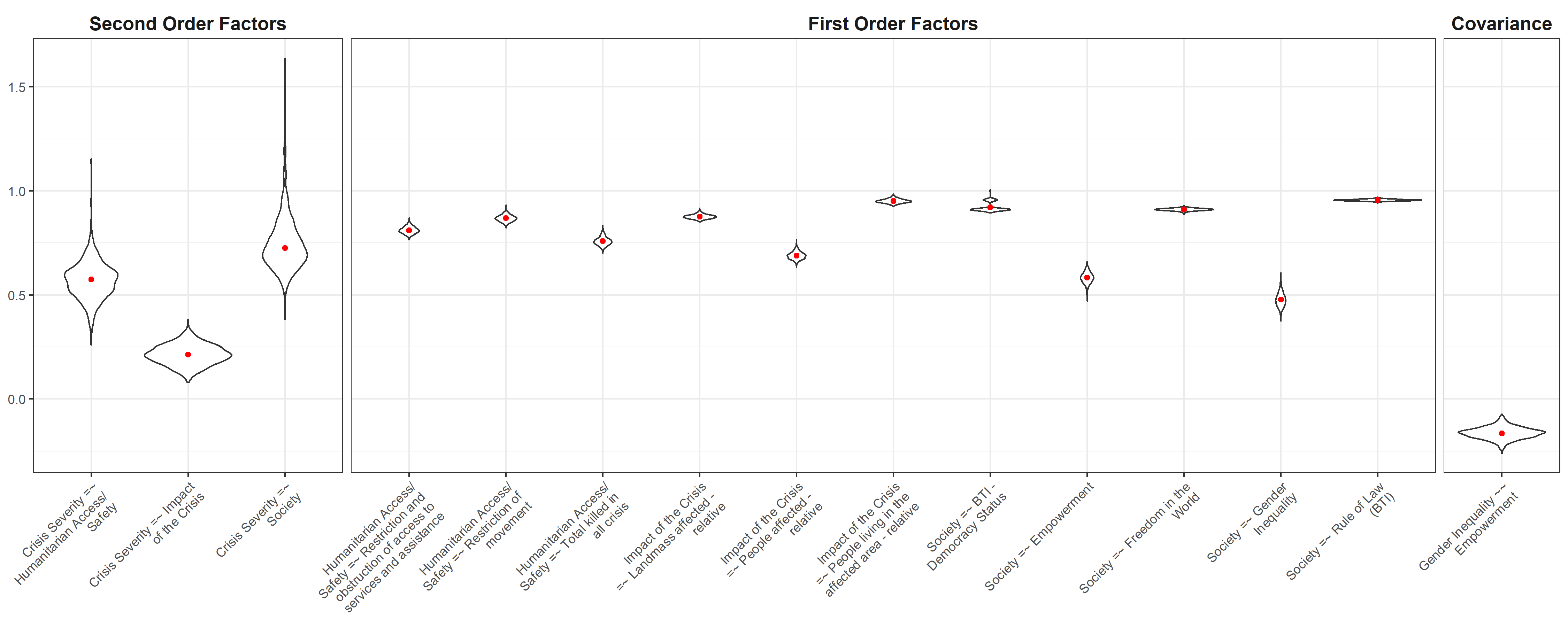

Appendix 10 Figure 1 shows comparable results to the CFA model fit presented in the main paper. Moreover, if the heuristics for acceptable fit are applied (CFI and TLI greater than 0.90 and RMSEA less than 0.08), all samples had acceptable CFI values and most samples had acceptable TLI values. RMSEA values, while slightly greater than the acceptable threshold, are inline with the values presented in the main paper. The slightly greater chi-squared estimate from the main paper CFA is likely due to the larger samples size used (i.e., larger degree of freedom).

Likewise, Appendix 10 Figure 2 also shows consistent results between the models run with partial datasets and the full dataset (that is, the main paper findings). The factor loadings from the main paper align with the mean of the samples in almost all cases.

Overall, the similarity between model fit statistics and factor loading values suggest that the analysis presented in the main paper is unlikely to be overfit to the data. Rather, this evidence suggests that the final CFA may perform consistently if new crises were added to the database.

1. Indicators: *Total People in Need* (referred to in the text as “People in Need”) and *Current humanitarian conditions of total population in the affected area* (referred to in the text as “People Affected”)
2. Muthén, L. K., & Muthén, B. O. (2002). How to use a Monte Carlo study to decide on sample size and determine power. *Structural equation modeling*, *9*(4), 599-620.
